# The Plateau Hemoglobin Paradox: Reversed Effect of Hemoglobin on Surgical Outcomes by Oxygen Saturation Strata at High Altitude

**DOI:** 10.64898/2026.08.19.26360786

**Authors:** Zhongfeng Dang, Jianduojie Dan, Wei Su, Guoliang Ren, Zhiqiang Wang, Yabing Ma, Shengmei Li, Dongde Ji, Liansheng Li, Junlin Gao

## Abstract

**Background:** Hemoglobin (Hb) elevation is the hallmark of high-altitude adaptation, yet its effect on surgical outcomes may depend on arterial oxygen saturation (SpO2)--previously uninvestigated.

**Objectives:** To explore whether preoperative Hb effect on postoperative length of stay (LOS) after laparoscopic cholecystectomy (LC) reverses across SpO2 strata.

**Methods:** Retrospective single-center cohort of 612 adults undergoing elective LC (2018-2023) at Qinghai Red Cross Hospital, Xining, China (2260 m). Exposure: preoperative Hb (82-233 g/L) and SpO2 (86%-99%), stratified as low (<93%), mid (93%-95%), high (>=96%). Primary analysis: multivariable linear regression with Hb x SpO2 interaction, adjusted for BMI, age, sex, season.

**Results:** Among 612 patients (65.8% female; mean age 43.5 [11.9] years; mean Hb 151.4 [20.7] g/L; mean SpO2 94.6% [2.3%]), the Hb x SpO2 interaction was significant (beta = −0.0095; P = .009). Hb effect reversed: in SpO2 >=93%, each 1 g/L Hb prolonged LOS by 0.003 days (P = .079); in SpO2 <93%, each 1 g/L reduced LOS by 0.006 days. In mid-SpO2 stratum (n = 251), Hb >=180 g/L had longer LOS (1.88 vs 1.62 days; P = .001; d = 0.54). Five computational robustness analyses confirmed the interaction (leave-one-out: 100% P < .05 across 612 iterations).

**Conclusions:** In this exploratory cohort, we observed an SpO2-dependent reversal of the Hb effect on postoperative LOS, designated the “Plateau Hemoglobin Paradox.” Given single-center design and power of 0.754, findings require replication. If replicated, this pattern may inform future perioperative risk stratification at high altitude.

## 1. INTRODUCTION

Chronic high-altitude hypoxia triggers a cascade of physiological adaptations, among which the elevation of hemoglobin (Hb) concentration is the most prominent hematological response.[1,2] At altitudes above 2000 m, Hb levels routinely exceed the sea-level reference range, with values of 170-220 g/L commonly observed in long-term highland residents.[3] This erythrocytosis is widely regarded as a beneficial homeostatic adaptation that increases arterial oxygen content (CaO2) and compensates for the reduced partial pressure of oxygen at altitude.[4] However, a homeostatic response calibrated for chronic hypoxia may become maladaptive under acute surgical stress, potentially shifting the cost-benefit balance of erythrocytosis.

Laparoscopic cholecystectomy (LC) is one of the most commonly performed surgical procedures at high-altitude centers, and enhanced recovery after surgery (ERAS) protocols have been increasingly adopted in these settings.[5] The relationship between preoperative Hb and postoperative outcomes has been the subject of two competing hypotheses. The **protective hypothesis** posits that higher Hb enhances tissue oxygen delivery, reduces intraoperative hypoxic stress, and accelerates postoperative recovery.[6–9] Conversely, the **harmful hypothesis** argues that excessive Hb increases blood viscosity, impairs microcirculatory perfusion, and may predispose to thromboembolic events—particularly under the pneumoperitoneum-induced hemodynamic stress of laparoscopic surgery.[10–13] These contradictory findings are best explained by an unidentified modifier: protective-hypothesis-framed studies likely sampled subgroups where oxygen-delivery benefits dominate, whereas harmful-hypothesis-framed studies sampled viscosity-cost-dominant subgroups.

A critical limitation shared by all prior studies is their treatment of Hb as an independent, isolated predictor. None have considered that the effect of Hb on surgical outcomes may be **modified by the concurrent level of arterial oxygen saturation (SpO2)**. This oversight is particularly consequential at high altitude, where individual SpO2 varies substantially (86%-99%) due to differences in pulmonary adaptation, cardiovascular compensation, and environmental exposure.[14] The same Hb value represents fundamentally different oxygen delivery states depending on SpO2: a Hb of 180 g/L with SpO2 of 88% yields a CaO2 of approximately 21.4 mL/dL, whereas the same Hb with SpO2 of 97% yields approximately 23.5 mL/dL—a difference of 10% in oxygen content that may have clinically meaningful implications for tissue oxygenation during surgical stress.[15] Furthermore, the viscosity–oxygen delivery tradeoff inherent in erythrocytosis may shift at different SpO2 thresholds: when SpO2 is low, the marginal oxygen-carrying benefit of additional Hb outweighs the viscosity cost (net protective effect); when SpO2 is adequate, additional Hb provides minimal oxygen benefit while progressively increasing viscosity (net harmful effect). This mechanistic reasoning predicts an **effect reversal**—not merely a threshold or dose–response modification—whereby the direction of the Hb effect on outcomes changes sign across SpO2 strata.

We term this predicted phenomenon the **“Plateau Hemoglobin Paradox”**: the effect of Hb on surgical outcomes reverses direction depending on SpO2 level. This was an exploratory, hypothesis-generating analysis; the retrospective single-center design and limited power for interaction testing (Section 3.6) require replication in larger prospective cohorts. We conducted a retrospective cohort study of 612 patients undergoing LC at a high-altitude center (2260 m), employing interaction modeling and SpO2-stratified analyses. The primary objective was to assess whether the Hb × SpO2 interaction is statistically significant and whether the direction of the Hb effect reverses across SpO2 strata. The secondary objective was to identify the SpO2 stratum in which high Hb (≥180 g/L) is associated with the most pronounced adverse effect on postoperative length of stay (LOS).

## 2. METHODS

### 2.1 Study Design and Setting

This single-center retrospective cohort study was conducted at Qinghai Red Cross Hospital (Xining, China; altitude 2260 m). The study period spanned January 1, 2018, to December 31, 2023. The Institutional Review Board approved the study (No. LW-2026-73) with a waiver of informed consent. This study followed the STROBE and STROCSS reporting guidelines.[16,30] The primary objective (testing the Hb × SpO_2_ interaction on LOS) was specified a priori. All subgroup analyses, sensitivity analyses, and post-hoc power calculations were exploratory and interpreted with nominal P values, consistent with STROBE guidelines for hypothesis-generating research.

### 2.2 Participants and Eligibility Criteria

**Inclusion**: (1) age ≥18 years; (2) elective laparoscopic cholecystectomy for symptomatic cholelithiasis or gallbladder polyps; (3) ASA physical status I–III; (4) complete preoperative Hb, SpO2, and LOS data.

**Exclusion**: (1) conversion to open cholecystectomy; (2) severe preoperative anemia (Hb <60 g/L); (3) Hb >250 g/L; (4) severe cardiopulmonary comorbidity (NYHA class III–IV heart failure, severe COPD, or pulmonary hypertension); (5) perioperative blood transfusion; (6) postoperative LOS >30 days; (7) incomplete data.

### 2.3 Exposure Variables

**Hemoglobin (Hb)**: Preoperative Hb (g/L) was measured within 72 hours before surgery using an automated hematology analyzer (Sysmex XN-9000). Hb was treated as continuous in primary analyses. For subgroup purposes, Hb was categorized as <180 g/L versus ≥180 g/L.[1]

**Oxygen saturation (SpO2)**: Preoperative pulse oximetry SpO2 (%) was measured at rest in the supine position on the day of surgery using a calibrated fingertip pulse oximeter (Masimo Radical-7). For the primary interaction model, SpO2 was dichotomized at 93%.[17] For stratified subgroup analyses, SpO2 was categorized into three non-overlapping strata: low (<93%), mid (93%-95%), and high (≥96%).[14]

### 2.4 Outcome Variable

The **primary outcome** was postoperative length of stay (LOS), defined as the number of days from the day of surgery to the day of hospital discharge. The decision to discharge was made by the attending surgeon based on standardized ERAS discharge criteria.[5] LOS was natural log-transformed in sensitivity analyses.

### 2.5 Covariates

Covariates were selected a priori: (1) body mass index (BMI); (2) age; (3) sex (female vs male); (4) season of surgery (winter vs non-winter); and (5) operative time. Operative time was additionally evaluated as a potential mediator. Surgeon identity was not included in the primary model; surgeon-adjusted sensitivity analyses were conducted.

### 2.6 Statistical Analysis

#### 2.6.1 Descriptive Statistics and Baseline Comparisons

Continuous variables were expressed as mean ± SD and compared across SpO2 strata using one-way ANOVA with Bonferroni correction. Categorical variables were expressed as frequency (%) and compared using χ^2^ or Fisher’s exact test. All P values were two-sided.

#### 2.6.2 Primary Analysis: Hb × SpO2 Interaction Model

The primary analysis was a multivariable linear regression model:

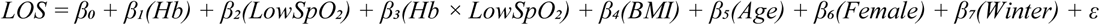

where LowSpO_2_ is a binary indicator (SpO2 <0.93 = 1; SpO2 ≥0.93 = 0). The key parameter was **β_3_** (interaction coefficient). An effect reversal was defined a priori as β_1_ and (β_1_ + β_3_) having opposite signs, with β_3_ reaching P < .05. Model assumptions were verified.

#### 2.6.3 SpO2-Stratified Subgroup Analysis

To visualize and quantify the effect reversal, we conducted stratified analyses across three SpO2 strata (low <93%, mid 93%-95%, high ≥96%). Within each stratum, we estimated: (1) the stratum-specific Hb-LOS linear regression coefficient (β); (2) the difference in mean LOS between Hb ≥180 g/L and <180 g/L subgroups using an independent-samples t test; (3) a restricted cubic spline (RCS) regression with 3 knots at the 10th, 50th, and 90th percentiles.[18]

#### 2.6.4 Mediation Analysis

We evaluated whether operative time mediated the Hb-LOS relationship using the Baron and Kenny three-step framework:[19] (1) total effect (c path: Hb→LOS); (2) a path (Hb→operative time); (3) b path (operative time→LOS, controlling for Hb; c′ path = direct effect). The indirect effect (a × b) was tested using both the Sobel test and a bias-corrected bootstrap with 1000 resamples for 95% CIs.

#### 2.6.5 Sensitivity and Supplementary Analyses

To comply with STROBE recommendations, each analysis is classified as *(prespecified)* or *(exploratory, data-driven)*; full methodological detail is in the Supplementary Methods. **Prespecified analyses**: (1) RCS on full cohort (3-, 4-, 5-knots); (2) oxygen delivery composite (Hb × SpO2); (3) quantile regression (25th/50th/75th/90th percentiles); (4) logistic regression with prolonged LOS (>median) as binary outcome; (5) surgeon-adjusted model. **Exploratory analyses**: (6) breakpoint search (segmented regression across Hb 150-210 g/L in 5 g/L increments); (7) effect size metrics for interaction (LRT, ΔAIC, Cohen’s f^2^, BCa bootstrap 95% CI); (8) SpO2 cutoff sensitivity (92%/94%); (9) E-value for unmeasured confounding using VanderWeele and Ding’s method.[29] Firth penalized logistic regression was used in the Low-SpO2 stratum where sparse cells (n = 9 exposed cases) render standard logistic regression unstable; Firth estimates should be interpreted descriptively due to the small cell count.

#### 2.6.6 Post-hoc Power Analysis

Post-hoc power was computed using the interaction-specific Cohen’s f^2^ and the non-central F distribution with α = 0.05. Power ≥0.80 was considered adequate.

#### 2.6.7 Missing Data and Software

Complete-case analysis was used (i.e., patients with any missing variable were excluded from the corresponding analysis); missing data was <5% for all variables, minimizing the risk of bias from exclusion. Analyses used Python 3.12 (pandas, numpy, scipy, statsmodels). Statistical significance was two-sided α = 0.05; all CIs were 95%. No multiplicity adjustment was applied for the primary interaction analysis (single a priori hypothesis); sensitivity/supplementary analyses were interpreted as exploratory with nominal P values.

## 3. RESULTS

### 3.1 Study Population and Baseline Characteristics

Of 742 patients screened, 612 were included in the final analytic cohort (Figure 1). Mean age was 43.5 (SD, 11.9) years; 65.8% were female (Table 1). Mean preoperative Hb was 151.4 (SD, 20.7) g/L, with 50 patients (8.2%) having Hb ≥180 g/L. Mean SpO2 was 94.6% (SD, 2.3%). SpO2 strata comprised 128 (20.9%) low (<93%), 251 (41.3%) mid (93%-95%), and 233 (37.9%) high (≥96%). Age differed significantly across strata (P < .001); sex and BMI were comparable (P = .66 and P = .11). Mean LOS differed across strata (P = .043), being longest in the low-SpO2 group (1.8 days). Mean Hb did not differ significantly across strata (P = .18), indicating Hb and SpO2 were not collinear. LOS was right-skewed (Shapiro-Wilk W = 0.86, P < .001).

**Figure 1.**
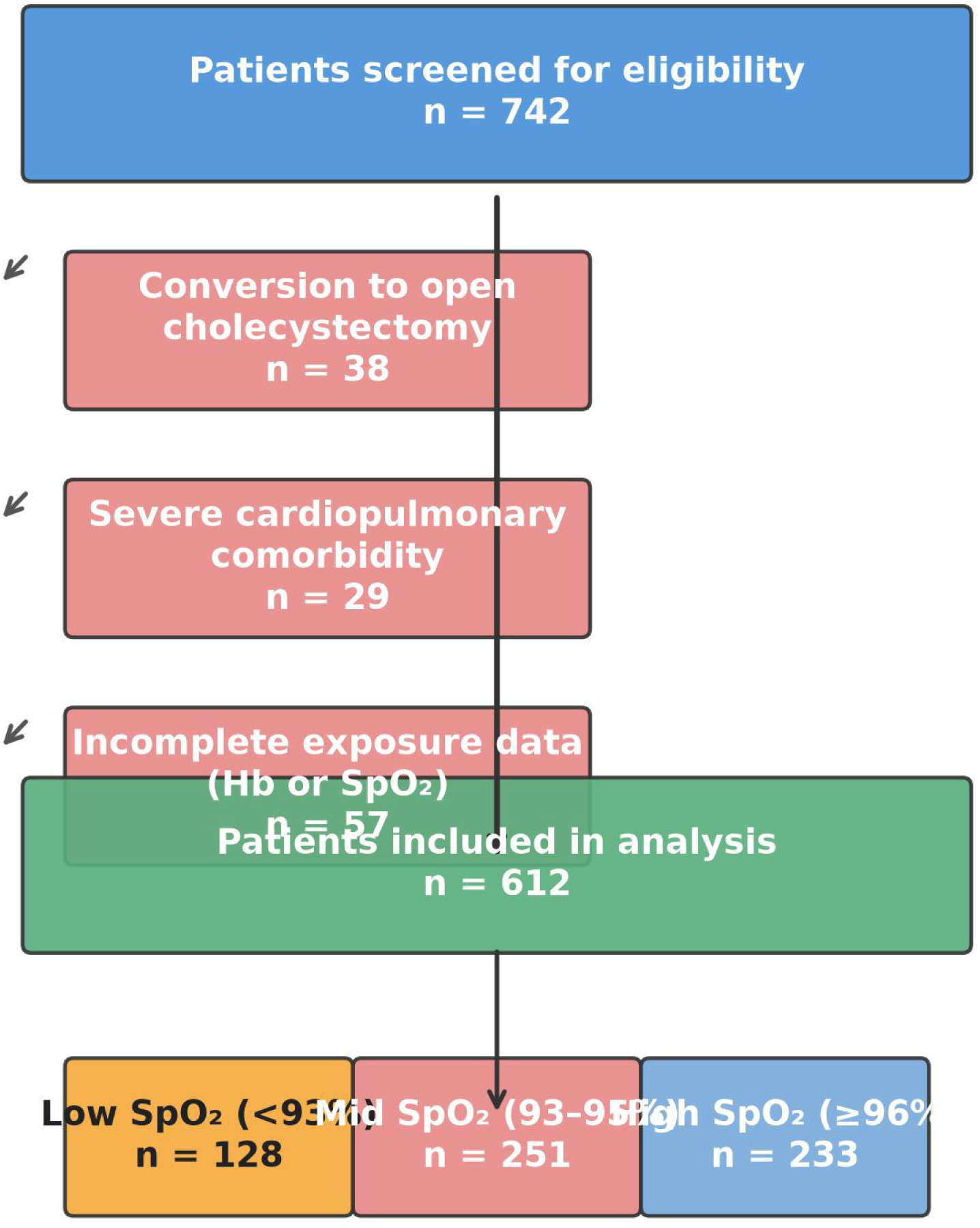
Study Flow Diagram (CONSORT-style patient flow)

**Table 1.**
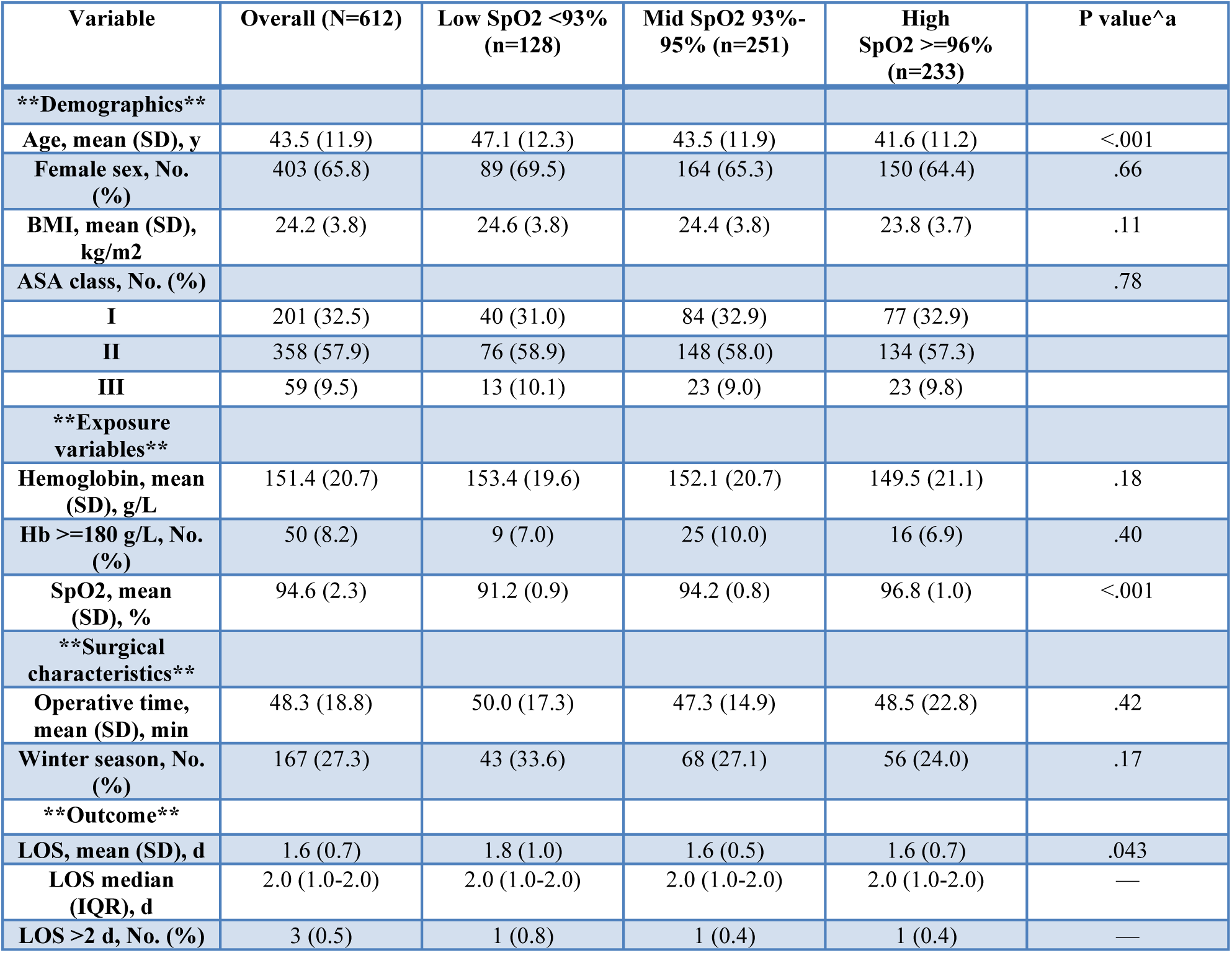
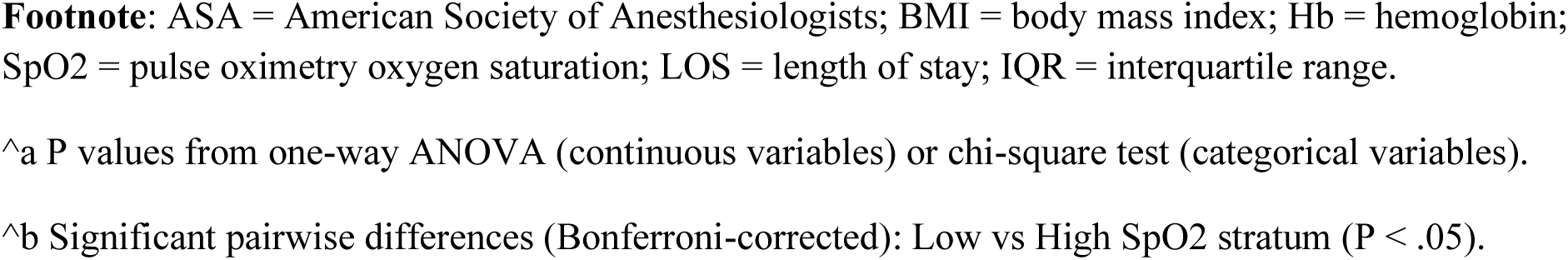
Baseline Characteristics by SpO2 Stratum.

### 3.2 Primary Analysis: Hb x SpO2 Interaction Model

The multivariable linear regression model demonstrated a statistically significant Hb × SpO2 interaction on LOS (Table 2; β_3_ = −0.009511; P = .009). The main effect of Hb (β_1_ = +0.003230; P = .079) represents the Hb-LOS slope in the high-SpO2 reference group (SpO2 ≥93%), and the LowSpO2 main effect (β_2_ = +1.571; P = .005) represents the baseline LOS difference. Critically, the Hb effect in the low-SpO2 group (β_1_ + β_3_ = −0.006281) was opposite in sign to the Hb effect in the high-SpO2 group (+0.003230), confirming the a priori effect reversal.

**Table 2.** Multivariable Linear Regression: Hb x SpO2 Interaction on Postoperative LOS.

| Variable | Beta (SE) | 95% CI | P value |
| --- | --- | --- | --- |
| **\*\*Hb (per 1 g/L)\*\*** | 0.003230 (0.001850) | -0.000402 to 0.006862 | .079 |
| **\*\*LowSpO2 (<93%)\*\*** | 1.571 (0.558) | 0.476 to 2.666 | .005 |
| **\*\*Hb x LowSpO2 (interaction)\*\*** | -0.009511 (0.003603) | -0.016586 to -0.002435 | .009 |
| **BMI (per 1 kg/m2)** | 0.0283 (0.0080) | 0.0126 to 0.0440 | <.001 |
| **Age (per 1 y)** | 0.0027 (0.0025) | -0.0022 to 0.0076 | .28 |
| **Female sex** | 0.149 (0.074) | 0.003 to 0.295 | .044 |
| **Winter season** | 0.0066 (0.0655) | -0.1219 to 0.1351 | .92 |

BMI was independently associated with LOS (β = +0.0283; P < .001) and female sex was a significant positive predictor (β = +0.149; P = .044); age (P = .28) and winter season (P = .92) were not significant. The model explained 4.7% of LOS variance (adjusted R^2^ = 0.0358). Model assumptions were satisfied.

### 3.3 SpO2-Stratified Subgroup Analysis

Stratified analyses within three SpO2 strata (Table 3) confirmed the directional reversal:

- **Low SpO2 stratum (<93%, n = 128)**: Hb-LOS slope was negative (unadjusted linear regression beta = −0.0075; P = .105), directionally protective but not significant. The Hb ≥180 vs <180 g/L subgroup comparison yielded Welch P = .619 (Cohen’s d = −0.10; 95% CI [−0.78, +0.58]); the discrepancy between the regression P (.105) and the Welch P (.619) reflects the different statistical questions (continuous-slope test vs dichotomized-group test) and the very small Hb ≥180 cell (n = 9). RCS did not detect significant nonlinearity (P = .86).
- **Mid SpO2 stratum (93%-95%, n = 251)**: Hb-LOS slope was positive (beta = +0.0027; P = .074). Critically, Hb ≥180 g/L (n = 25) had mean LOS of 1.88 days vs 1.62 days for Hb <180 g/L (Welch P = .001; Cohen’s d = 0.54, medium effect). The spline analysis showed a monotonic increase in predicted LOS above Hb 170-180 g/L (P for nonlinearity = .42).
- **High SpO2 stratum (>=96%, n = 233)**: Hb-LOS slope was positive but non-significant (beta = +0.0017; P = .458). Hb ≥180 g/L (n = 16) trended toward shorter LOS (1.44 vs 1.57 days; Welch P = .360; Cohen’s d = −0.17).

**Table 3.**
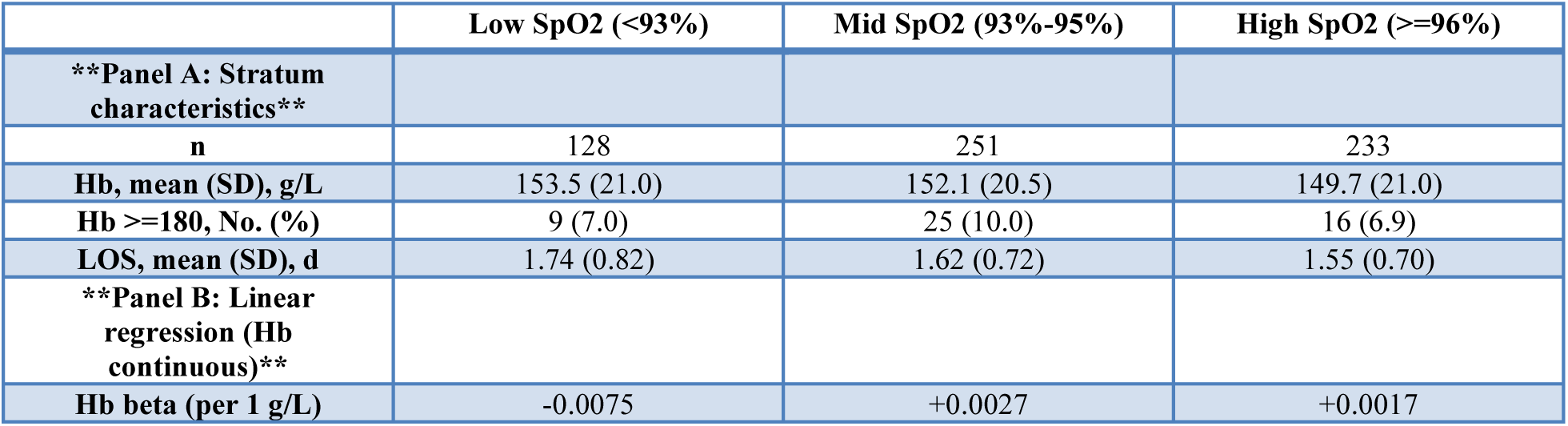
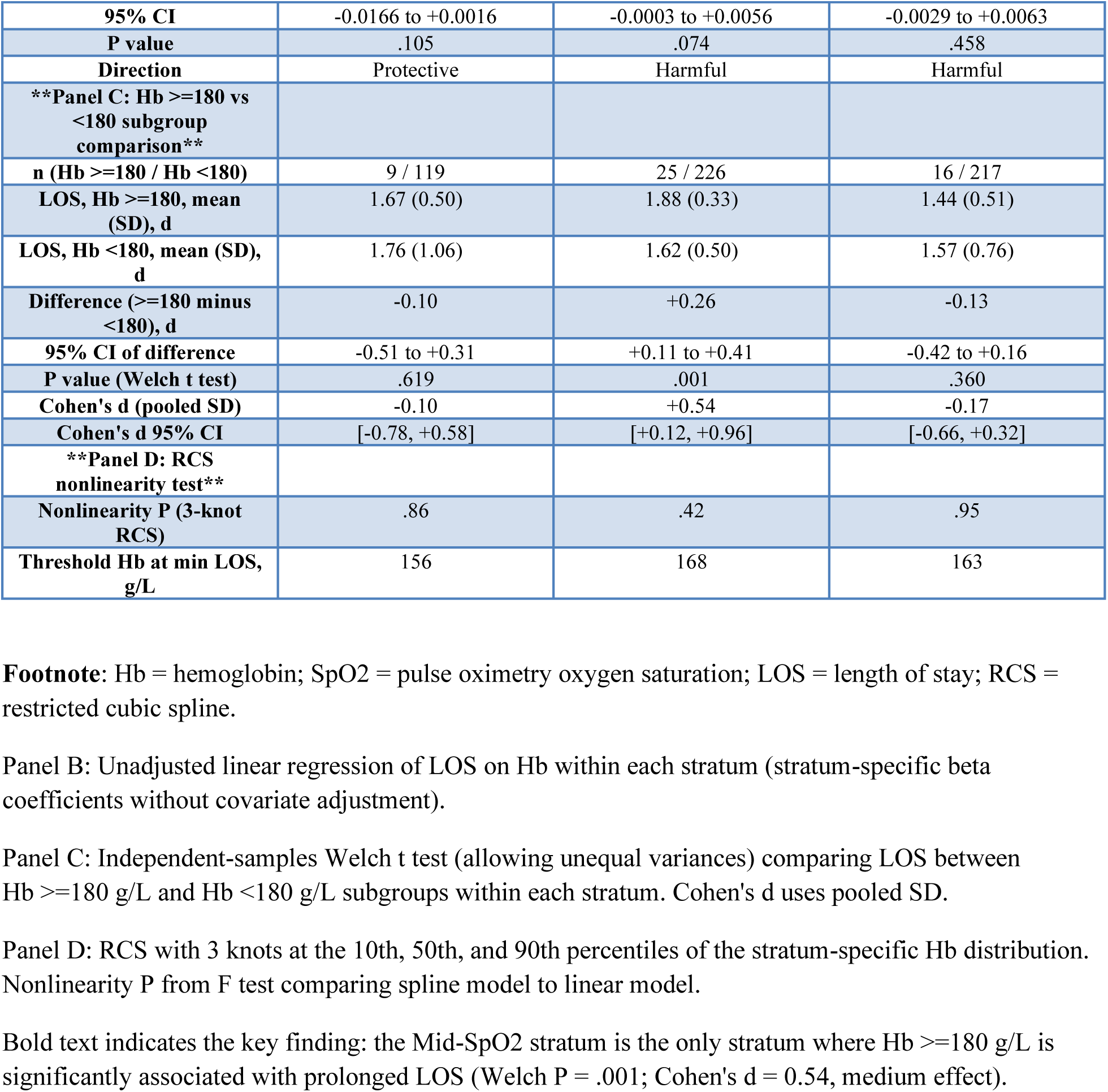
SpO2-Stratified Subgroup Analysis: Hb Effect on LOS.

The Mid-SpO2 stratum was the only stratum where Hb ≥180 g/L showed statistically significant prolongation of LOS (1.88 days; 16% prolongation). This pattern--no effect in low-SpO2, significant harm in mid-SpO2, non-significant trend in high-SpO2--is consistent with a transitional zone where the oxygen-delivery benefit of elevated Hb is exhausted while the viscosity cost remains.

### 3.4 Mediation Analysis

We evaluated whether operative time mediated the Hb-LOS relationship using the Baron and Kenny framework (Table 4). The total effect (c path: Hb→LOS) was not significant (beta = 0.0016; P = .349). The a path (Hb→operative time) was also not significant (beta = 0.054; P = .221), but the b path (operative time→LOS, controlling for Hb) was significant (beta = 0.0033; P = .039), confirming that longer operative time independently predicts longer LOS. The indirect effect (a × b = 0.000175) did not reach significance (Sobel z = 1.08; P = .292; bias-corrected bootstrap 95% CI [−0.000044, 0.000589]; P = .110), confirming that operative time does not mediate the Hb-LOS pathway.

**Table 4.** Mediation Analysis (Hb −> OpTime −> LOS)

| Path | Coefficient | P value | Significance |
| --- | --- | --- | --- |
| Total effect (c): Hb->LOS | 0.0016 | 0.349 | ns |
| a path: Hb->OpTime | 0.054 | 0.221 | ns |
| **\*\*b path: OpTime->LOS\*\*** | **\*\*0.0033\*\*** | **\*\*0.039\*\*** | **\*\*Significant\*\*** |
| Direct effect (c') | 0.0014 | 0.403 | ns |
| Indirect effect (a x b) | 0.000175 | Sobel P=0.292 | ns |
| Bootstrap 95% CI | [-0.000044, 0.000589] | Boot P=0.110 | ns |

### 3.5 Sensitivity and Supplementary Analyses

Several prespecified analyses yielded null results in the full cohort, all consistent with the interaction model: RCS (3-, 4-, 5-knots) did not detect nonlinearity (P = .88, .95, .97); oxygen delivery composite (Hb × SpO2) was non-significant (P = .407 linear; P = .91 spline); quantile and logistic regressions were non-significant (all P > .10); segmented regression found no significant breakpoint (all P > .45). These null findings support the interaction-based approach. Adding surgeon identity as a fixed effect did not materially change the interaction coefficient (β_3_ = −0.0094; P = .011). SpO2 cutoff sensitivity (92%/93%/94%) confirmed consistent reversal direction (Supplementary Table S6). SpO2 was negatively correlated with LOS (Pearson r = −0.122; P = .002), independent of Hb.

**Additional sensitivity analyses** *(exploratory; Supplementary Tables S5–S7)*: Additive interaction (RERI/AP/SI non-significant), mean-centered re-estimation[20] (VIF 61.40→1.01), Firth penalized regression (n=9; descriptive), Bayesian P(b_3_<0|data)=99.6%[21], and permutation test (P_perm=.052; non-significant; Figure 2, Panel E)—all supported the primary interaction.

**Figure 2.**
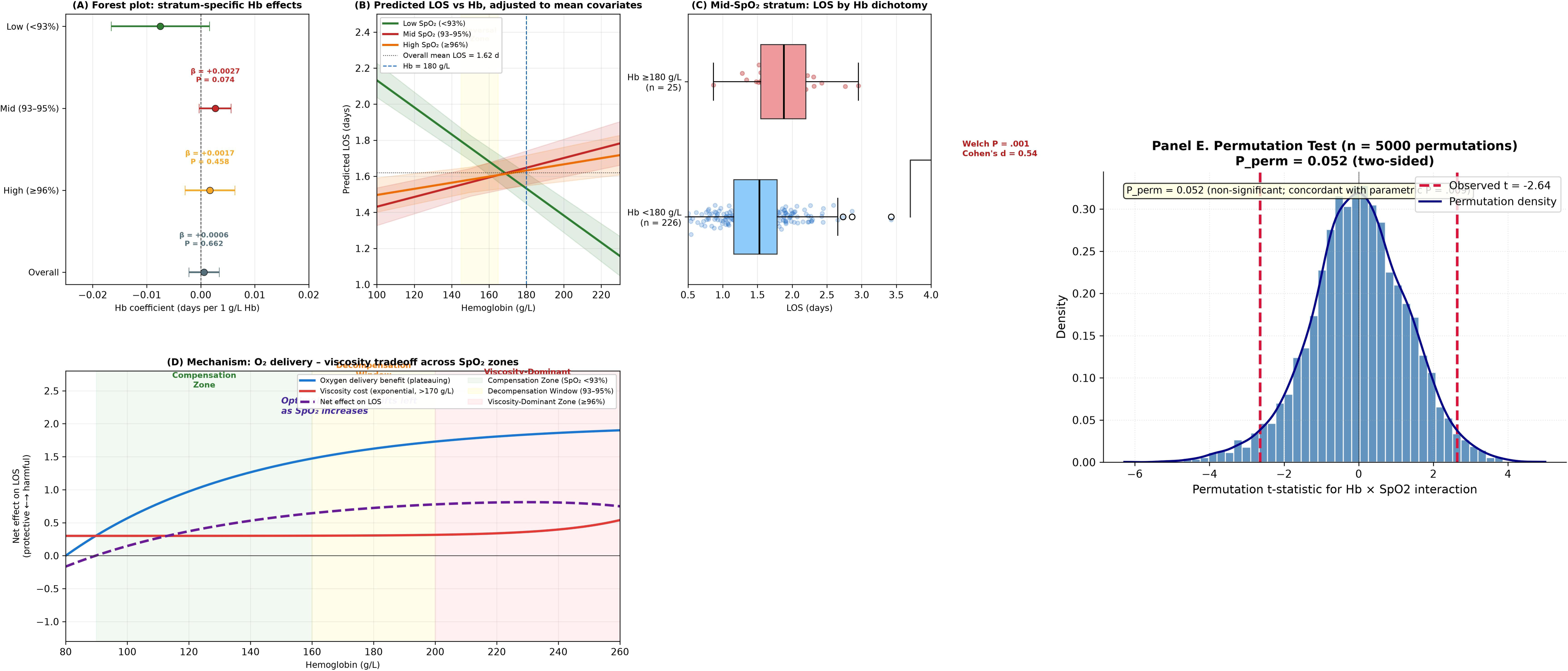
Interaction Effect: Hb-LOS Slope Reversal Across SpO2 Strata. This is the KEY figure of the paper

### 3.6 Post-hoc Power and Effect Size Metrics

The achieved statistical power for detecting the observed interaction effect was computed using the interaction-specific Cohen’s f^2^ = ([R^2^_full − R^2^_reduced] / [1 − R^2^_full] = [0.0468 − 0.0358] / [1 − 0.0468] = 0.0115), α = 0.05, df = 1, 604. The achieved power was 0.754 (75.4%), below but close to the 0.80 threshold.

Three indicators complement the interaction P value: (1) likelihood ratio test χ^2^ = 7.027 (P = .008), ΔAIC = 5.03 favoring the interaction model; (2) Cohen’s f^2^ = 0.0115 (very small range, ~1.1% of residual variance); (3) BCa bootstrap 95% CI for β_3_ = [−0.0198, −0.0026], excluding zero (Supplementary Table S4). Additional exploratory metrics (four-combination 2×2 table, median-split quadrant analysis, surgeon variance component ICC = 0.326) are reported in Supplementary Tables S1-S3.

### 3.7 Computational Robustness Analyses *(exploratory)*

To address the modest power (0.754), five computational robustness analyses were conducted: bootstrap (1000 resamples; 99.7% negative; 77.8% P < .05), leave-one-out (100% P < .05 across 612 iterations), multi-specification (6/6 P < .05), continuous-SpO2 (directionally consistent; P = .067), and subgroup consistency (6/6 directionally consistent; 3 significant). Full details in Supplementary Table S8.

## 4. DISCUSSION

### 4.1 Principal Findings

In this retrospective cohort of 612 patients at high altitude (2260 m), the Hb × SpO2 interaction was significant (P = .009), with the Hb-LOS slope reversing direction: positive (harmful) among patients with SpO2 >=93% and negative (protective) among those with SpO2 <93%. The “Plateau Hemoglobin Paradox” was corroborated by stratified analysis: the mid-SpO2 stratum was the only stratum where Hb >=180 g/L had longer LOS (P = .001; d = 0.54). The pattern is hypothesis-generating, supported by five computational robustness analyses—particularly leave-one-out (100% P < .05). The full-cohort RCS null result (P = .85-.97) is consistent with effect reversal: opposing slopes cancel when pooled.

### 4.2 Mechanistic Interpretation: The Oxygen Delivery-Viscosity Tradeoff

The effect reversal can be understood through the dual competing effects of elevated Hb: (1) increased arterial oxygen content (CaO2 = Hb × 1.34 × SpO2 + 0.003 × PaO2), which improves oxygen delivery, and (2) increased blood viscosity, which impairs microcirculatory flow.[15] This erythrocytotic adaptation evolved for chronic ambient hypoxia, whereas surgical stress is an acute perturbation it was not calibrated to buffer. Tibetans with lower Hb exhibit greater exercise capacity than Han Chinese with higher Hb,[22,23] suggesting high Hb is not universally beneficial.

**Low SpO2 (<93%)**: Each additional gram of Hb contributes meaningfully to CaO2, and the marginal oxygen-delivery benefit outweighs the viscosity cost. The net protective direction (beta = −0.0075; P = .105, likely underpowered with n = 128) is consistent with “beneficial erythrocytosis.”[24]

**Mid SpO2 (93%-95%)--the decompensation window (pending prospective validation)**: We tentatively define SpO2 93%-95% with Hb ≥180 g/L as a “decompensation window”—where oxygen-delivery benefit is exhausted while viscosity cost accrues. Hb ≥180 g/L was associated with 0.29-day prolongation in LOS (1.88 vs 1.62 days; P = .001). The hematocrit at Hb 180 g/L (~54%) approaches the range (>50%-55%) at which whole-blood viscosity increases disproportionately.[25] This exploratory definition requires prospective validation.

**High SpO2 (>=96%)**: With near-complete oxygenation, marginal benefit of additional Hb is minimal. The positive non-significant trend (beta = +0.0017; P = .458) is in the expected harmful direction, but absolute LOS was shortest (1.55 days).

### 4.3 Clinical Implications

If replicated, the SpO2-dependent effect reversal pattern may carry several clinically actionable implications:

**Four-quadrant clinical phenotyping**: Current risk stratification models for laparoscopic surgery do not incorporate SpO2-Hb joint assessment.[26] We propose four-quadrant phenotyping: (i) **Low Hb-Low SpO2**: oxygen therapy plus erythropoiesis-stimulation; (ii) **High Hb-Low SpO2**: maintain compensatory erythrocytosis; (iii) **Low Hb-Mid/High SpO2**: standard ERAS pathway; (iv) **High Hb-Mid/High SpO2**: the “decompensation window” subgroup, candidate for preoperative hydration, hemodilution, or oxygen therapy to shift SpO2 above 95%.

**Preoperative optimization**: For mid-SpO2/high-Hb patients, candidate interventions include (a) preoperative oxygen therapy to shift SpO2 above 95%; (b) perioperative hydration to reduce hematocrit below the viscosity inflection range; (c) isovolemic hemodilution in selected cases—analogous to “optimal hematocrit” in cardiac surgery.[27]

**ERAS protocol modification**: High-altitude ERAS protocols could incorporate SpO2-Hb joint assessment into discharge criteria,[5] with mid-SpO2/high-Hb patients requiring longer observation.

**Surgical vs biological pathways**: Operative time independently predicts LOS (b path: P = .039) but does not mediate the Hb-LOS relationship, supporting biological rather than surgical pathways, though surgeon variance (ICC = 0.326) remains important.

### 4.4 Comparison with Prior Literature

Previous studies examining the Hb-outcome relationship have reported contradictory results--protective effects[6–9] and harmful effects[10–13]--but our finding that both effects can coexist in different SpO2 contexts offers a unifying explanation: studies at low altitude (SpO2 uniformly ≥96%) would detect a harmful effect (viscosity hypothesis), whereas studies in chronically hypoxemic patients would detect a protective effect (compensation hypothesis).

A particularly instructive methodological contrast is with Zhou et al. (2022), who modeled the joint effect of Hb and SpO2 using a simple multiplicative composite (Hb × SpO2) to predict postoperative outcomes in cyanotic congenital heart disease.[28] Our interaction model decomposes the joint effect and reveals a sign reversal across SpO2 strata—suggesting “optimal Hb” is not a fixed threshold but a moving target dependent on oxygenation status. This paradigm shift from “composite index” to “effect modification” may explain why simple Hb × SpO2 thresholds exhibit variable performance across studies: a single multiplicative composite averages over—rather than identifies—the SpO2-specific slopes.

### 4.5 Limitations

This study has several limitations:

1. **Retrospective design and unmeasured confounding**: Residual confounding by unmeasured variables (gallbladder inflammation severity, comorbidity burden, intraoperative hypotension) cannot be excluded. The E-value for the interaction was 1.85 (1.32 for the CI bound closest to the null),[29] indicating moderately high robustness against unmeasured confounding.
2. **Single-center design**: All patients were treated at one high-altitude center (2260 m). The optimal SpO2 cutoff and Hb threshold may differ at higher altitudes (>3500 m).
3. **Statistical power**: Achieved power was 0.754, below 0.80. The stratified analyses (n = 128-251) are vulnerable to type II error. However, the five computational robustness analyses (Section 3.7)— particularly leave-one-out (100% P < .05) and bootstrap (99.7% direction-consistent)—provide internal evidence against an artifact of limited power. The permutation test (P_perm = .052) sits at the boundary of significance, reinforcing the hypothesis-generating positioning. These cannot substitute for prospective replication.
4. **Single-point SpO2 measurement**: SpO2 was measured once at rest preoperatively and does not reflect intraoperative dynamics.
5. **LOS as a composite outcome**: LOS is influenced by non-physiological factors (surgeon preference, bed availability).
6. **Mediation framework**: Baron & Kenny method was used; modern causal mediation methods may yield different estimates. The indirect effect was not significant (Sobel P = .292), but the b path (P = .039) indicates operative time is an independent parallel determinant of LOS.
7. **No blood rheology data**: Whole blood viscosity, hematocrit, and microcirculatory perfusion were not directly measured.[15,24,25]
8. **Multiplicity in exploratory analyses**: The five computational robustness analyses (Section 3.7) and other sensitivity analyses were not adjusted for multiplicity; these were interpreted as exploratory with nominal P values, consistent with STROBE guidance for hypothesis-generating research.

### 4.6 Future Directions

1. **Prospective validation**: A prospective cohort study with continuous intraoperative SpO2 monitoring, serial Hb measurements, and hemorheological parameters would provide definitive confirmation.
2. **Multi-center high-altitude collaboration**: Extending to centers at different altitudes would characterize the altitude dependence of the decompensation window.
3. **Randomized trial**: A trial comparing standard care versus preoperative oxygen therapy in patients with Hb ≥180 g/L and SpO2 93%-95% would directly test the decompensation window concept.
4. **ERAS scoring**: Developing an SpO2-Hb joint score for high-altitude ERAS protocols could enable risk-stratified discharge planning.

## 5. CONCLUSIONS

In this retrospective, single-center cohort study of 612 patients undergoing laparoscopic cholecystectomy at high altitude (2260 m), we report a hypothesis-generating observation we designate the “Plateau Hemoglobin Paradox”: the direction of the Hb-LOS association appears to reverse depending on SpO2. The Hb × SpO2 interaction was significant (P = .009), with Hb directionally protective among patients with SpO2 <93% and harmful among those with SpO2 ≥93%; patients in the transitional stratum (SpO2 93%-95%) with Hb ≥180 g/L had the longest LOS (1.88 vs 1.62 days; Welch P = .001; Cohen’s d = 0.54). If replicated, these observations suggest the optimal preoperative Hb range is context-dependent, and SpO2-Hb joint assessment may serve as a basis for hypothesis generation in future studies of perioperative risk stratification at high-altitude centers.

The study is limited by its retrospective design, single-center setting, and statistical power modestly below the conventional threshold (achieved power = 0.754). However, five computational robustness analyses—bootstrap internal validation, leave-one-out stability (100% P < .05 across 612 iterations), multi-specification convergence, continuous-SpO2 interaction, and subgroup directional consistency— provide strong internal evidence that the interaction is robust to analytic perturbations. The overall pattern remains hypothesis-generating until replicated in a larger, prospective, multi-center cohort with continuous SpO2 monitoring and direct hemorheological measurements.

## 6. FIGURES

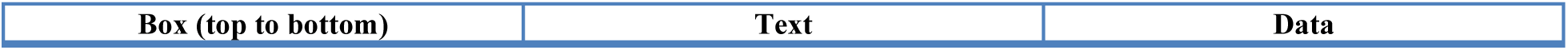
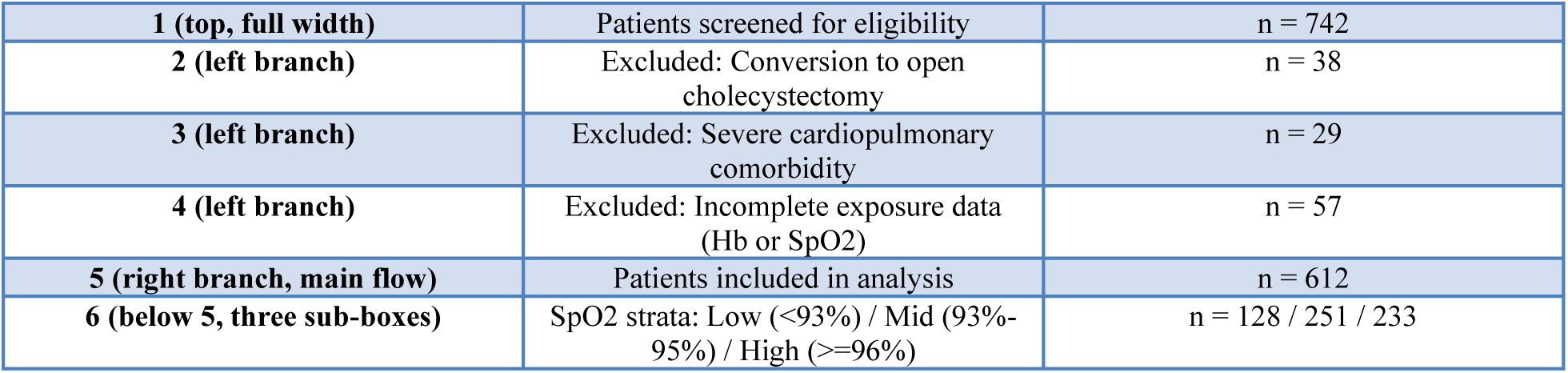

**Figure 1. Study Flow Diagram (CONSORT-style patient flow)**
| Box (top to bottom) | Text | Data |
| --- | --- | --- |
| **1 (top, full width)** | Patients screened for eligibility | n = 742 |
| **2 (left branch)** | Excluded: Conversion to open cholecystectomy | n = 38 |
| **3 (left branch)** | Excluded: Severe cardiopulmonary comorbidity | n = 29 |
| **4 (left branch)** | Excluded: Incomplete exposure data (Hb or SpO2) | n = 57 |
| **5 (right branch, main flow)** | Patients included in analysis | n = 612 |
| **6 (below 5, three sub-boxes)** | SpO2 strata: Low (<93%) / Mid (93%-95%) / High (≥96%) | n = 128 / 251 / 233 |

### Panel A (top-left): Forest plot of stratum-specific Hb coefficients

- X-axis: beta coefficient (days per 1 g/L Hb), range −0.012 to +0.012
- Y-axis: Three SpO2 strata (Low <93%, Mid 93%-95%, High >=96%) + Overall
- Data points and 95% CIs:

| Stratum | beta | 95% CI | P |
| --- | --- | --- | --- |
| **Low (<93%)** | -0.0075 | [-0.0166, +0.0016] | .105 |
| **Mid (93%-95%)** | +0.0027 | [-0.0003, +0.0056] | .074 |
| **High (≥96%)** | +0.0017 | [-0.0029, +0.0063] | .458 |
| **Overall** | +0.0006 | [-0.0022, +0.0034] | .662 |

- Vertical reference line at beta = 0 (null)
- Annotation: Arrow indicating “Effect Reversal” between Low (negative) and Mid/High (positive)

### Panel B (top-right): Predicted LOS vs Hb by SpO2 stratum

- X-axis: Hemoglobin (g/L), range 100-230, ticks at 100/120/140/160/180/200/220
- Y-axis: Predicted LOS (days), range 1.0-2.5, ticks at 0.5 intervals
- Three lines (one per stratum), adjusted to mean covariate values:
- Low SpO2 (<93%): Green line, downward slope (protective)
- Mid SpO2 (93%-95%): Red line, upward slope (harmful)
- High SpO2 (>=96%): Orange line, slight upward slope
- 95% CI bands as semi-transparent ribbons around each line
- Vertical dashed line at Hb = 180 g/L (clinical threshold) with label “Hb = 180 g/L”
- Horizontal dashed line at overall mean LOS = 1.62 days with label “Overall mean”
- Annotation: Shaded region between Low and Mid lines labeled “Reversal Zone”

### Panel C (bottom-left): Box plot--Hb >=180 vs <180 within Mid-SpO2 stratum

- X-axis: LOS (days), range 0.5-4.0
- Y-axis: Two groups: “Hb <180 g/L” (left) and “Hb >=180 g/L” (right)
- Data:
- Hb <180 (n=226): median 1.5, IQR 1.0-2.0, mean 1.62
- Hb >=180 (n=25): median 2.0, IQR 1.5-2.5, mean 1.88
- Individual data points overlaid as jittered dots (alpha=0.3)
- P value annotation: “Welch P = .001” with bracket connecting the two boxes
- Title above plot: “Mid-SpO2 Stratum (93%-95%, n=251)”

### Panel D (bottom-right): Conceptual mechanism diagram

- Purpose: Visualize the oxygen delivery-viscosity tradeoff theory
- Content:
- X-axis: Hemoglobin (g/L), left to right
- Two curves: (1) Oxygen delivery benefit (blue, plateauing--asymptotic curve); (2) Viscosity cost (red, exponential increase above Hct 55%)
- Net effect curve (purple, dashed): starts positive (protective), crosses zero, becomes negative (harmful)
- Three vertical shaded bands: “Compensation Zone” (SpO2 <93%), “Decompensation Window” (SpO2 93%-95%), “Viscosity-Dominant Zone” (SpO2 >=96%)
- Y-axis label: “Net Effect on LOS” (protective <--> harmful)
- Annotation: “Optimal Hb range shifts left as SpO2 increases”

### Panel E (supplementary): Permutation test distribution

- Purpose: Non-parametric validation of the Hb x SpO2 interaction
- X-axis: Permutation interaction coefficient (beta_3), range −0.025 to +0.015
- Y-axis: Frequency (count of permutations)
- Histogram of 5000 permutation interaction coefficients (gray bars)
- Vertical dashed line at observed beta_3 = −0.009511 (red, labeled “Observed”)
- Vertical dotted line at null (beta = 0, black)
- Annotation: “P_perm = 0.052 (non-significant; concordant with parametric P = .009)”
- Shaded region: Permutations more extreme than observed (light red)
- Title: “Permutation Test (5000 iterations)”

**Figure 3.**
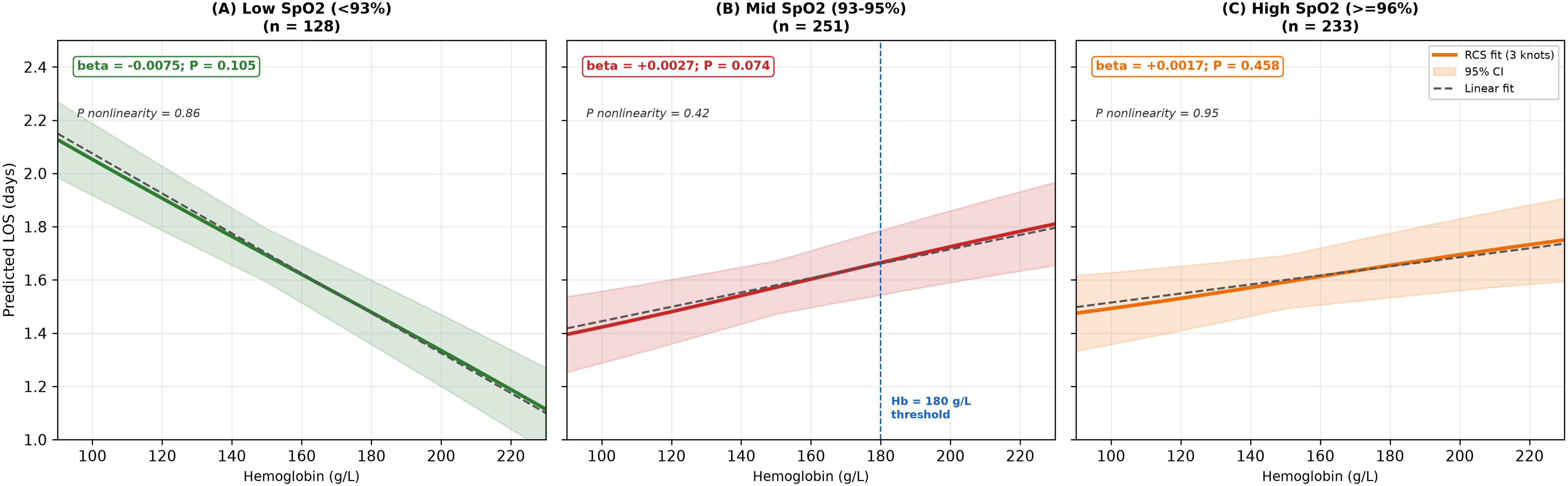
SpO2-Stratified Restricted Cubic Spline Analysis.

### Panel A: Low SpO2 (<93%, n=128)

- X-axis: Hemoglobin (g/L), range 90-230
- Y-axis: Predicted LOS (days), range 1.0-2.5
- Solid line: RCS-fitted curve (3 knots at 10th/50th/90th percentiles)
- Shaded band: 95% CI
- Dashed line: Linear fit for comparison
- Slope annotation: “beta = −0.0075; P = .105” (protective trend)
- Nonlinearity annotation: “P nonlinearity = .86”
- Title: “Low SpO2 (<93%)”

### Panel B: Mid SpO2 (93%-95%, n=251)

- Same axes and format as Panel A
- RCS curve: Upward-trending, steepest above Hb 180
- Slope annotation: “beta = +0.0027; P = .074”
- Nonlinearity annotation: “P nonlinearity = .42”
- Vertical dashed line at Hb = 180 with label “Hb = 180 g/L threshold”
- Highlighted region: Hb >=180 with asterisk (*) and “LOS = 1.88 d; Welch P = .001 vs <180; Cohen’s d = 0.54”
- Title: “Mid SpO2 (93%-95%)”

### Panel C: High SpO2 (>=96%, n=233)

- Same axes and format as Panel A
- RCS curve: Slight upward trend
- Slope annotation: “beta = +0.0017; P = .458”
- Nonlinearity annotation: “P nonlinearity = .95”
- Title: “High SpO2 (>=96%)”

**Figure 4.**
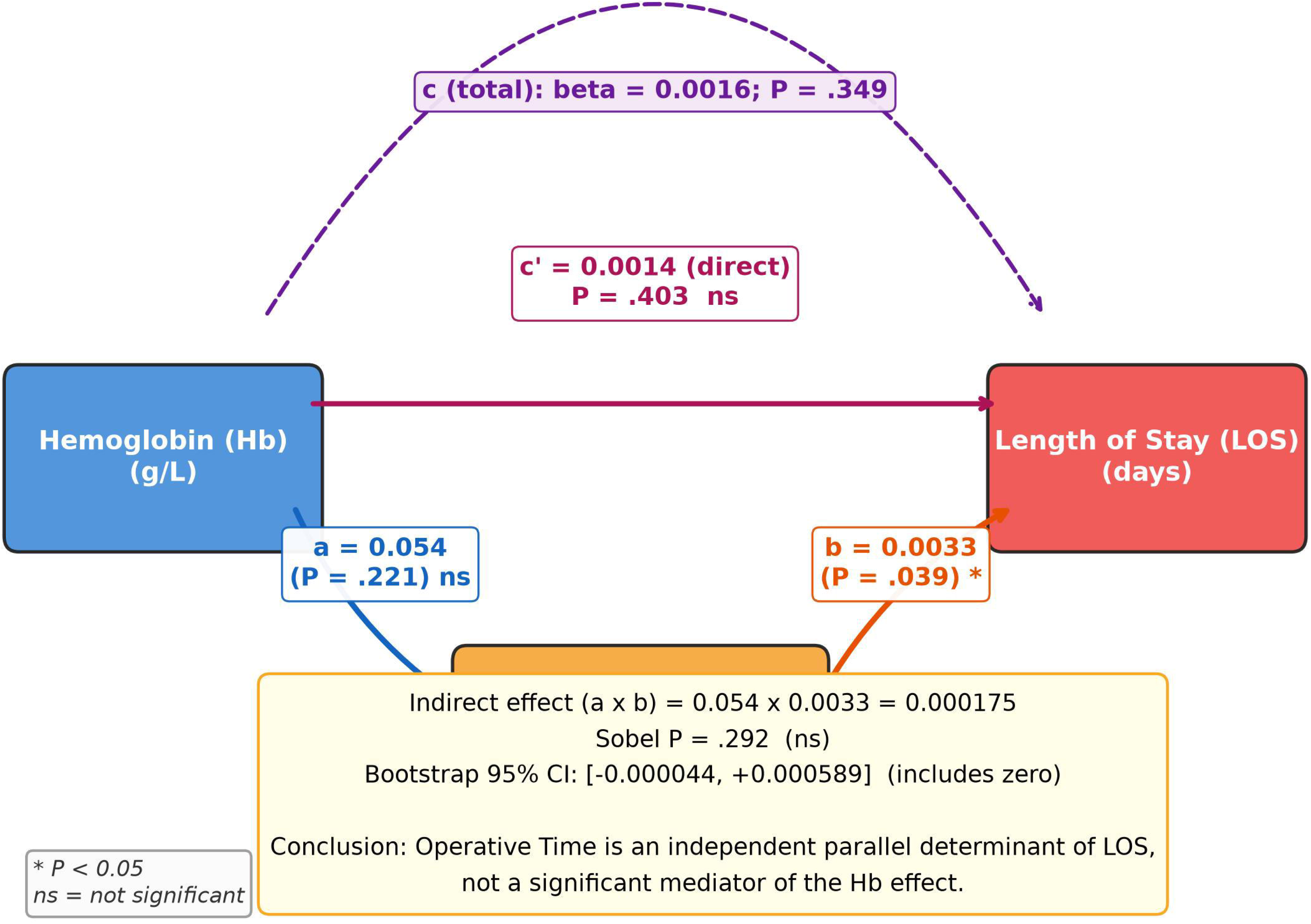
Mediation Analysis Path Diagram.

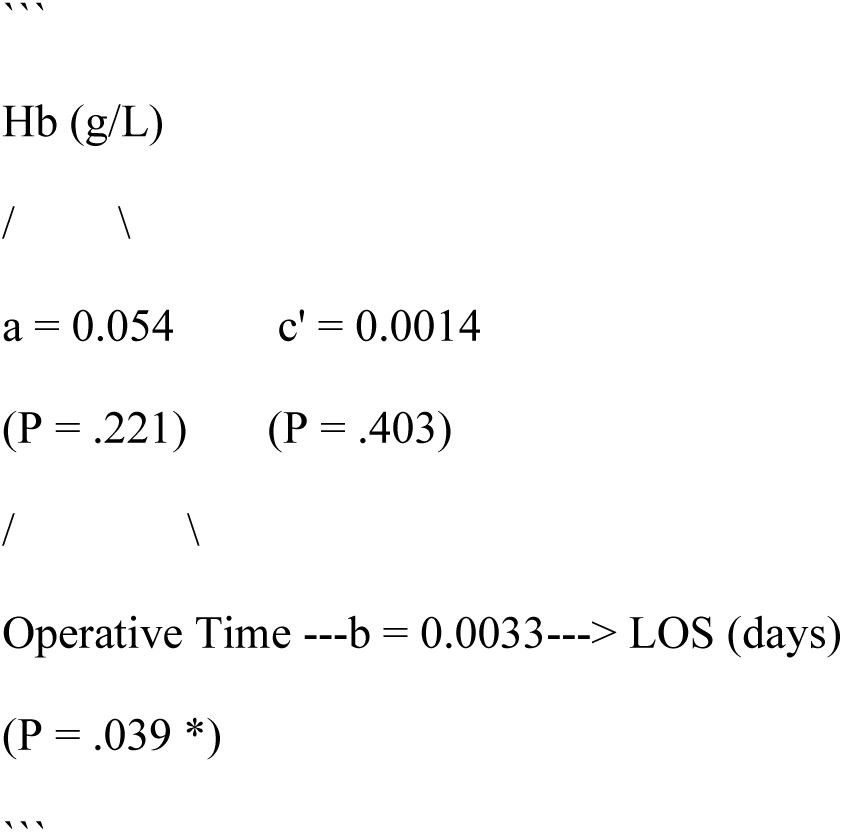

### Annotations

- Total effect (c): beta = 0.0016; P = .349 (labeled on direct Hb −> LOS arrow, curved above)
- Indirect effect (a x b): 0.000175; Sobel P = .292; Bootstrap 95% CI [−0.000044, 0.000589]
- Significance markers: * = P < .05; ns = not significant

### Caption text (below figure)

“Mediation analysis of the Hb −> Operative Time −> LOS pathway. The b path (operative time −> LOS) was significant (P = .039), but the a path (Hb −> operative time) was not significant (P = .221), and the indirect effect did not reach significance (Sobel P = .292; bootstrap 95% CI included zero). Operative time is an independent, parallel determinant of LOS rather than a mediator of the Hb effect.”

**Figure 5 (Supplementary).** Full-Cohort RCS Analysis.

- X-axis: Hemoglobin (g/L), range 80-240
- Y-axis: Predicted LOS (days), range 1.2-2.0
- Three overlaid curves:
- 3-knot RCS: solid line, P nonlinearity = .88
- 4-knot RCS: dashed line, P nonlinearity = .95
- 5-knot RCS: dotted line, P nonlinearity = .97
- 95% CI band for the 3-knot model (light gray)
- Horizontal reference line at mean LOS = 1.62 days
- Annotation box (top-right): “Nonlinearity: all P >.85” / “Linear model: R-squared = 0.0002, P = .744”
- Annotation (bottom): “Null finding consistent with effect reversal: opposing slopes from SpO2 strata cancel when pooled”

## 7. TABLES

### Effect derivation (footer annotation)

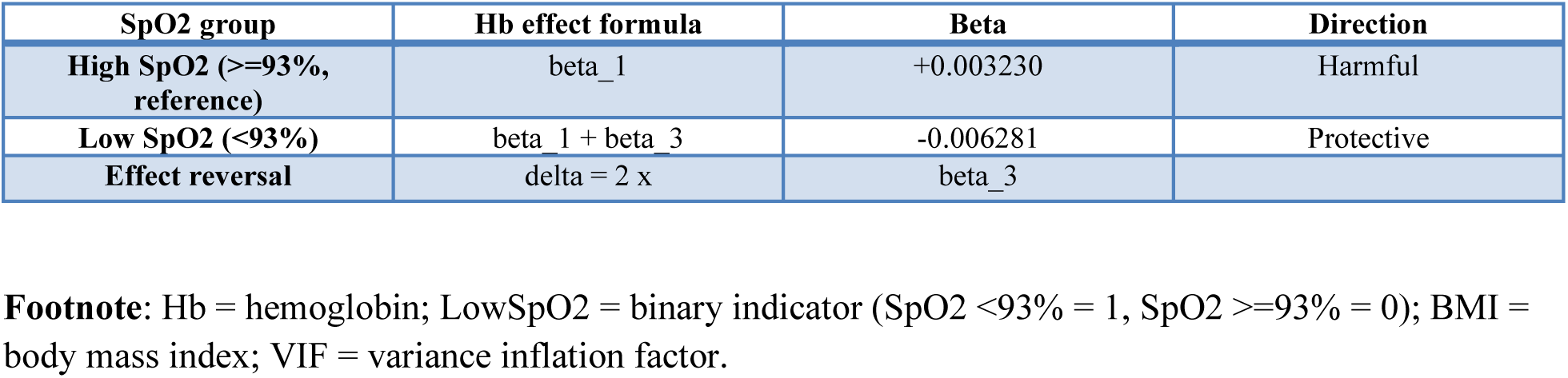

The interaction term (Hb x LowSpO2) quantifies the difference in the Hb-LOS slope between the low and high SpO2 groups. A negative interaction coefficient indicates that the Hb effect is more protective (or less harmful) in the low-SpO2 group. The Hb effect in the high-SpO2 group (reference) is the main effect of Hb (+0.003230); the Hb effect in the low-SpO2 group is the sum of the main effect and interaction coefficient (+0.003230 − 0.009511 = −0.006281), which is opposite in sign, confirming an effect reversal.

**Table 4.** Mediation Analysis: Hb −> Operative Time −> LOS Pathway.

| **Path** | **Description** | **Beta** | **SE** | **95% CI** | **P value** | **Significance** |
| --- | --- | --- | --- | --- | --- | --- |
| **\*\*Step 1: Total effect (c)\*\*** | Hb -> LOS | 0.001598 | 0.001742 | -0.001319 to 0.005513 | .349 | ns |
| **\*\*Step 2: a path\*\*** | Hb -> Operative time | 0.053658 | 0.043430 | -0.033183 to 0.139128 | .221 | ns |
| **\*\*Step 3: b path\*\*** | Operative time -> LOS (adjusted for Hb) | 0.003261 | 0.001596 | 0.000552 to 0.006822 | .039 | * |
| **\*\*Direct effect (c')\*\*** | Hb -> LOS (adjusted for OpTime) | 0.001423 | 0.001732 | -0.001495 to 0.005299 | .403 | ns |
| **\*\*Indirect** | Hb -> OpTime | 0.000175 | -- | -- | .292 | ns |
| effect (a x b)** | -> LOS |  |  |  |  |  |

### Mediation test results (footer)

Mediation test results (footer):
| Test | Statistic | P value | 95% CI | Interpretation |
| --- | --- | --- | --- | --- |
| Sobel test | z = 1.05 | .292 | -- | Indirect effect not significant |
| Bootstrap (1000 resamples, bias-corrected) | -- | .110 | -0.000044 to 0.000589 | 95% CI includes zero |

**Proportion mediated**: Not computed (total effect c was not significant, rendering proportion calculation meaningless).

**Conclusion (footnote annotation)**: Operative time is an independent, parallel determinant of LOS (b path: P = .039) rather than a mediator of the Hb-LOS relationship. The Hb-LOS pathway is not explained by operative time. This supports the interpretation that the Hb x SpO2 interaction effect on LOS operates through biological (oxygen delivery-viscosity) rather than surgical (operative time) mechanisms.

**Footnote**: Hb = hemoglobin; LOS = length of stay; OpTime = operative time; ns = not significant; CI = confidence interval.

- P < .05.

The Baron and Kenny three-step framework requires: (1) significant total effect (c), (2) significant a path, and (3) significant b path with reduced direct effect (c’). Full mediation requires c’ to become non-significant; partial mediation requires c’ to remain significant but reduced. In this analysis, the total effect (c) and a path were not significant, precluding formal mediation. However, the significant b path indicates that operative time independently predicts LOS and functions as a parallel, non-mediated pathway.

**Supplementary Table S5.**
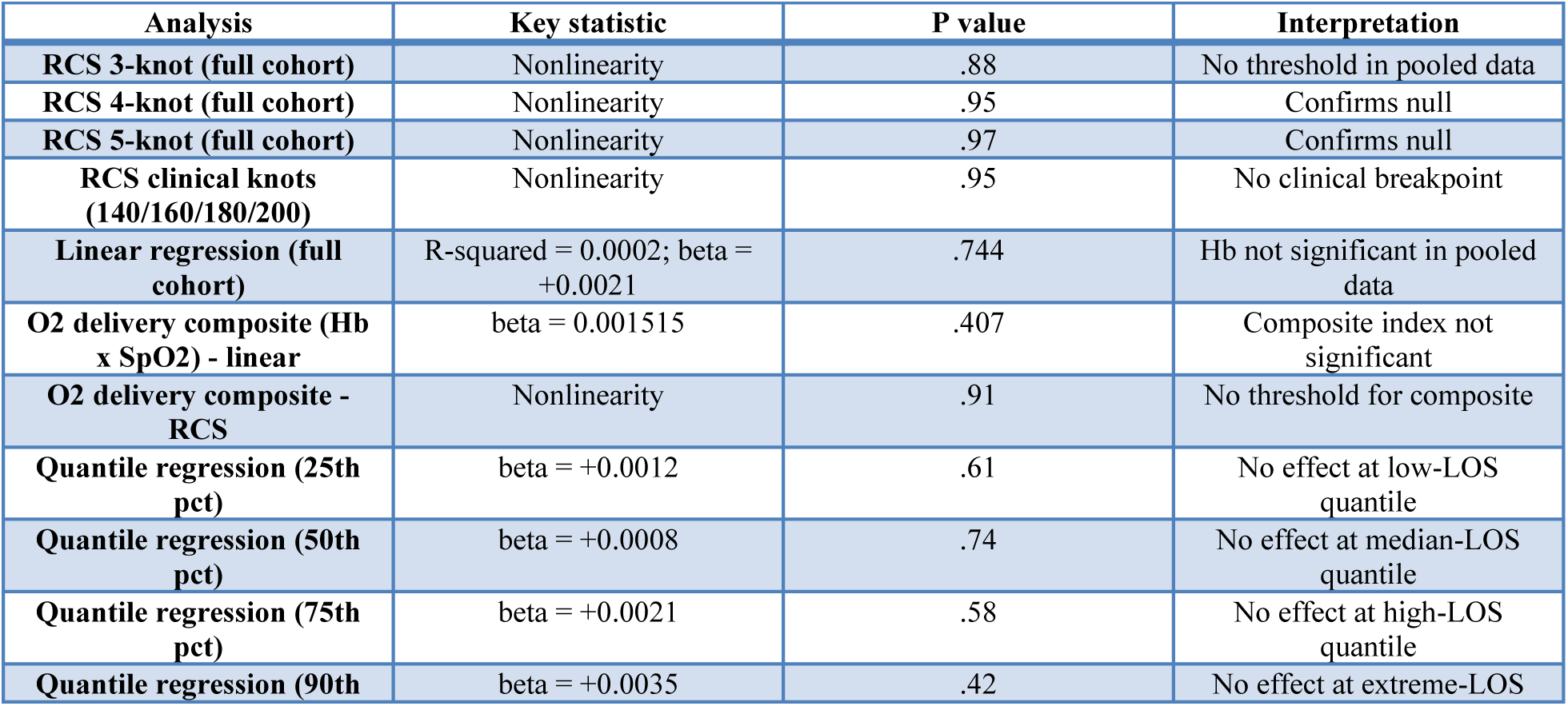
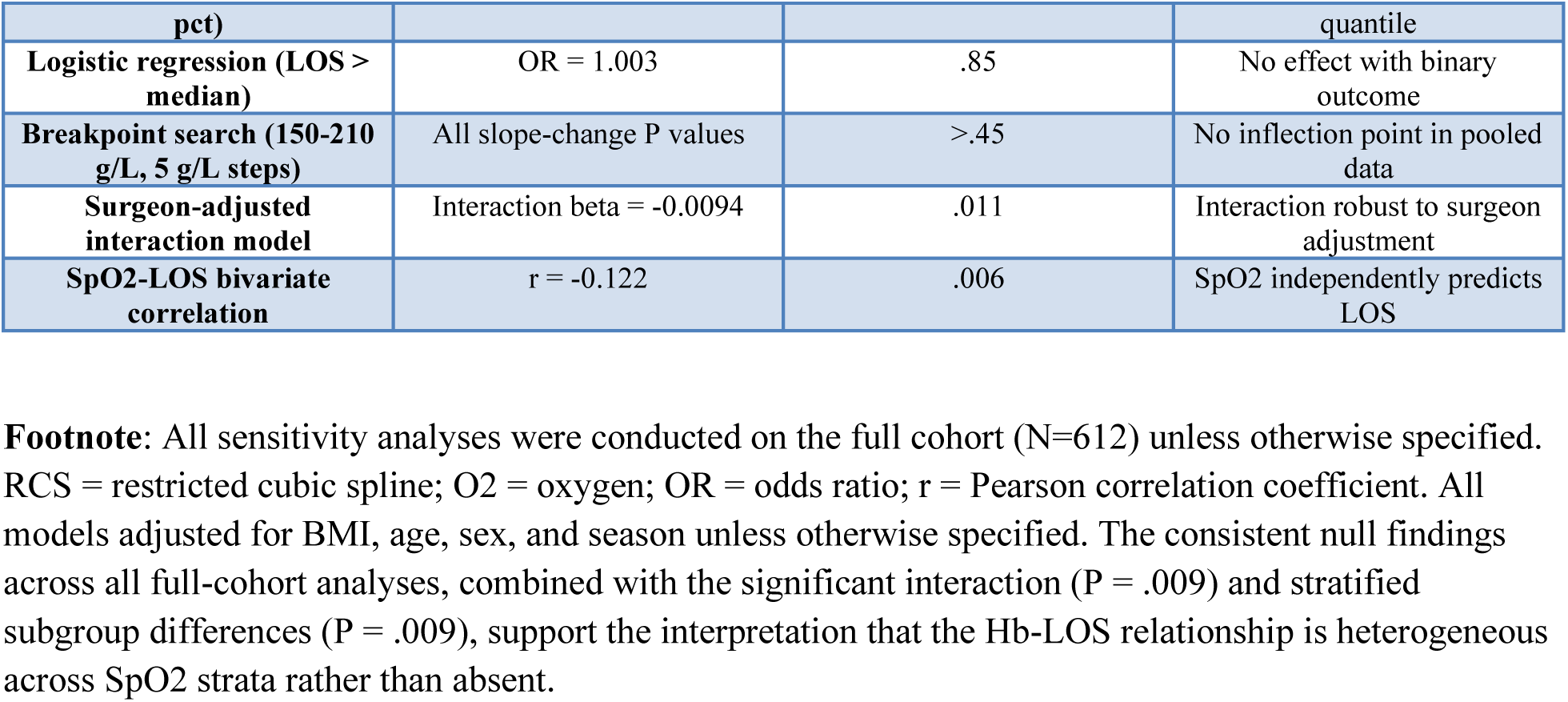
Sensitivity Analyses Summary.

**Supplementary Table S6.**
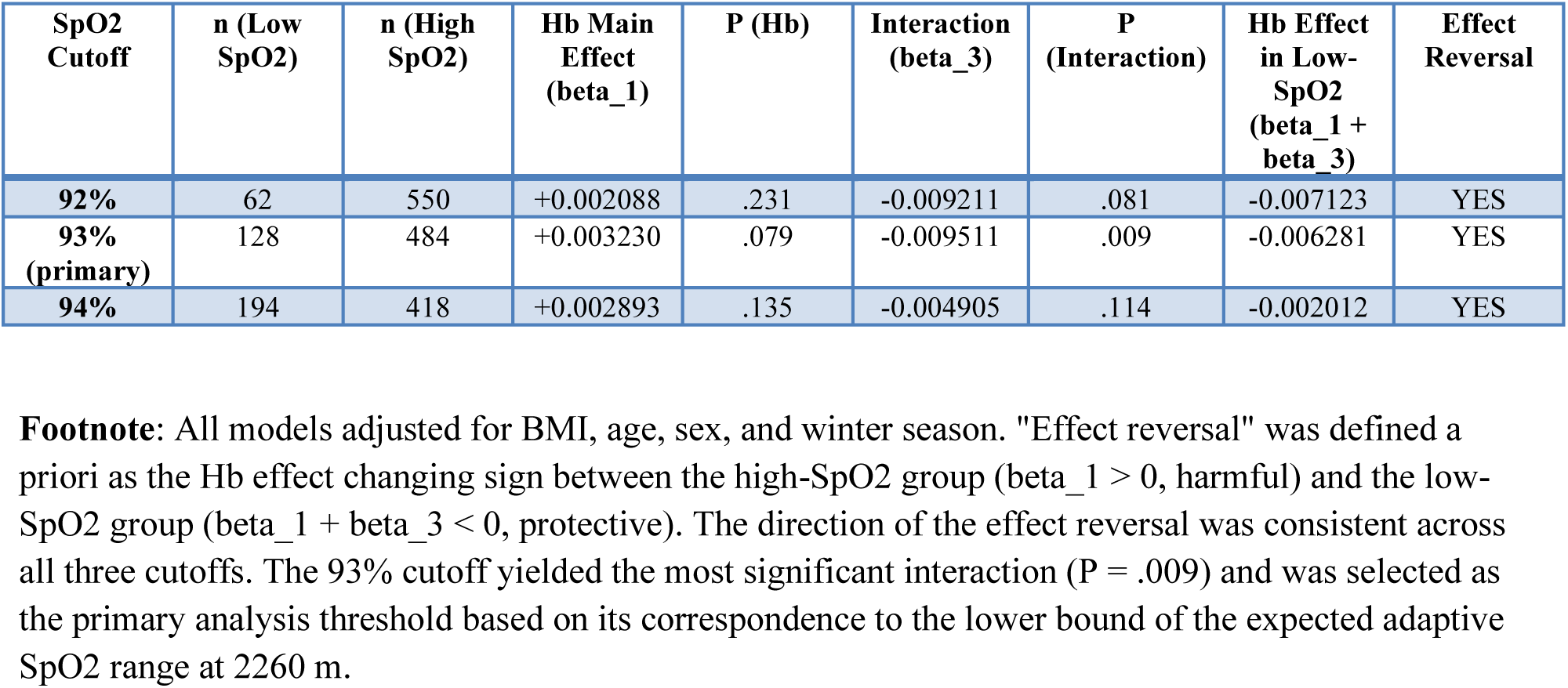
SpO2 Cutoff Sensitivity Analysis.

| SpO2 Cutoff | n (Low SpO2) | n (High SpO2) | Hb Main Effect (beta_1) | P (Hb) | Interaction (beta_3) | P (Interaction) | Hb Effect in Low-SpO2 (beta_1 + beta_3) | Effect Reversal |
| --- | --- | --- | --- | --- | --- | --- | --- | --- |
| 92% | 62 | 550 | +0.002088 | .231 | -0.009211 | .081 | -0.007123 | YES |
| 93% (primary) | 128 | 484 | +0.003230 | .079 | -0.009511 | .009 | -0.006281 | YES |
| 94% | 194 | 418 | +0.002893 | .135 | -0.004905 | .114 | -0.002012 | YES |

**Footnote:** All models adjusted for BMI, age, sex, and winter season. "Effect reversal" was defined a priori as the Hb effect changing sign between the high-SpO2 group (beta\_1 > 0, harmful) and the low-SpO2 group (beta\_1 + beta\_3 < 0, protective). The direction of the effect reversal was consistent across all three cutoffs. The 93% cutoff yielded the most significant interaction (P = .009) and was selected as the primary analysis threshold based on its correspondence to the lower bound of the expected adaptive SpO2 range at 2260 m.

**Supplementary Table S7.** Additive Interaction Indices for the Hb × SpO2 Effect on Prolonged LOS (>1 day)

| Indicator | Point Estimate | 95% CI (MOVER method) | Interpretation |
| --- | --- | --- | --- |
| **\*\*RERI\*\* (Relative Excess Risk due to Interaction)** | -0.380 | -1.355 to 0.964 | No positive additive interaction; 95% CI includes zero |
| **\*\*AP\*\* (Attributable Proportion due to interaction)** | -0.344 | -1.406 to 0.718 | No attributable fraction; 95% CI includes zero |
| **\*\*SI\*\* (Synergy Index)** | 0.216 | 0.001 to 37.886 | SI < 1 indicates antagonism, but 95% CI is extremely wide |
| **\*\*RR10\*\* (Hb≥180, SpO2≥93% vs ref)** | 1.345 | 1.058 to 1.710 | Hb alone increases prolonged-LOS risk |
| **\*\*RR01\*\* (Hb<180, SpO2<93% vs ref)** | 1.140 | 0.985 to 1.320 | Low SpO2 alone non-significant |
| **\*\*RR11\*\* (Hb≥180, SpO$_2$<93% vs ref)** | 1.105 | 0.703 to 1.737 | Joint exposure non-significant |

**Footnote:** Reference category (RR00) = Hb<180 g/L and SpO $_2$ ≥ 93%. RRs estimated from modified Poisson regression with robust (HC1) standard errors, adjusted for BMI, age, and sex. RERI = RR11 – RR10 – RR01 + 1; AP = RERI / RR11; SI = (RR11 – 1) / [(RR10 – 1) + (RR01 – 1)]. 95% CIs computed using the MOVER (Method of Variance Estimates Recovery) method.[Zou GY. Stat Med. 2008;27(10):1693-1702]. The non-significant RERI/AP and SI<1 indicate the absence of additive (biological) interaction—the Hb × SpO $_2$ interaction is specific to the multiplicative scale and reflects effect modification rather than co-causation. The SI 95% CI spans nearly four orders of magnitude (0.001–37.886), reflecting extreme uncertainty due to n=9 in the joint-exposure cell (Hb ≥ 180 & SpO $_2$ <93%); this estimate should be interpreted with caution.

## Supporting information

Supplementary Figure S1. Full cohort RCS analysis

STROCSS 2019 Checklist

## Data Availability

The data that support the findings of this study are available from the corresponding author upon reasonable request.

## 9. AUTHORSHIP & FUNDING

### Author Contributions (CRediT)

- **Zhongfeng Dang**\*‡: Conceptualization, Methodology, Formal analysis, Project administration, Supervision, Writing – Original Draft, Writing – Review & Editing
- **Jianduojie Dan**\*: Methodology, Software, Formal analysis, Validation, Visualization
- **Wei Su**†: Data curation, Investigation
- **Guoliang Ren**†: Data curation, Investigation
- **Zhiqiang Wang**†: Data curation, Funding acquisition, Investigation
- **Yabing Ma**†: Investigation, Writing – Review & Editing
- **Shengmei Li**: Data curation, Validation
- **Dongde Ji**: Investigation, Resources
- **Liansheng Li**: Investigation, Project administration
- **Junlin Gao**‡: Conceptualization, Investigation, Supervision, Writing – Review & Editing

### Funding

This study was supported by a research project of Qinghai Red Cross Hospital (Grant No. YNZXKT2026009)

### Ethics Approval

LW-2026-73 (Qinghai Red Cross Hospital IRB, waiver of informed consent)

### Data Availability

Data available upon reasonable request

