## Supplementary figures and images for "The Plateau Hemoglobin Paradox: Reversed Effect of Hemoglobin on Surgical Outcomes by Oxygen Saturation Strata at High Altitude"

### Supplementary Figure S1. Full cohort RCS analysis

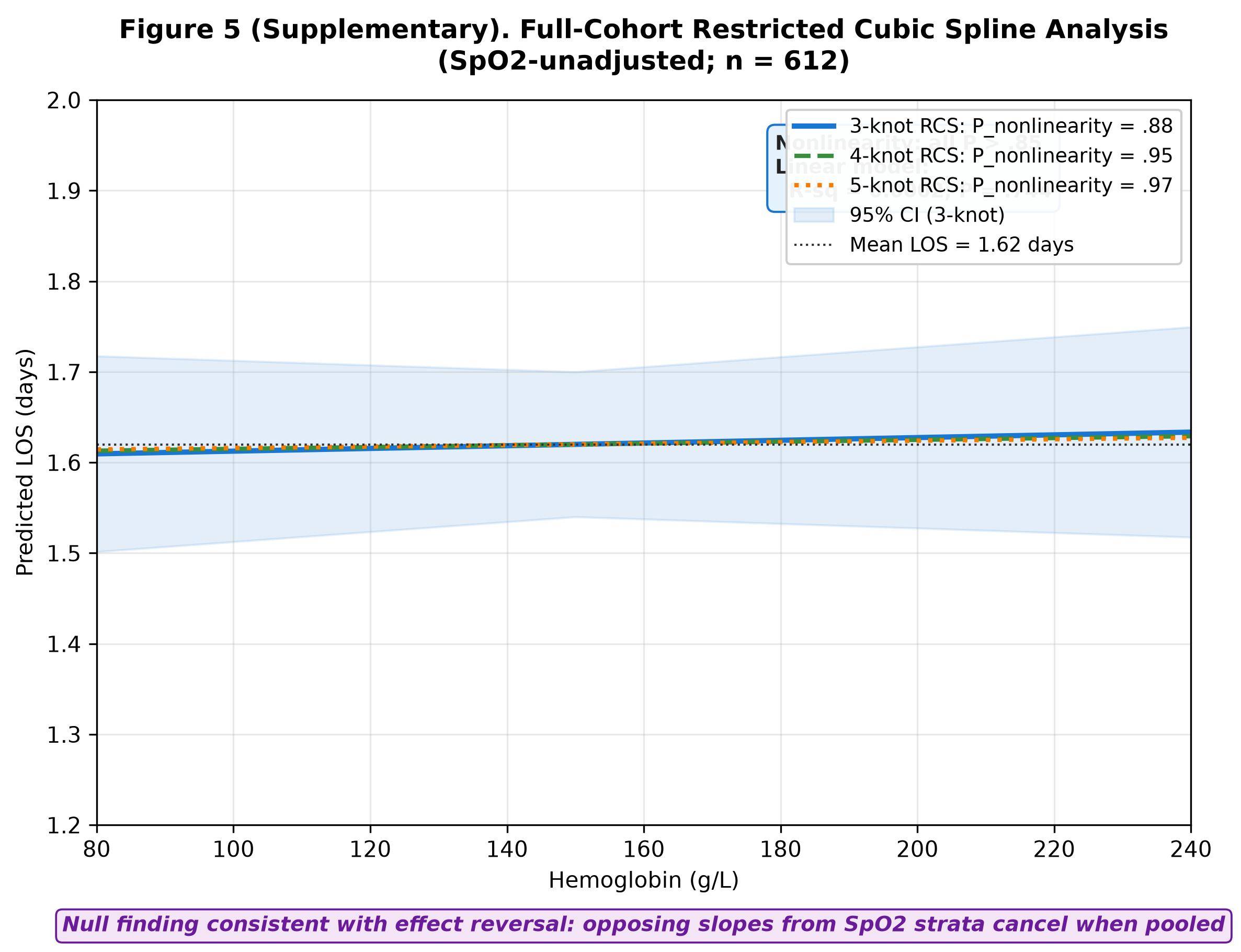
