## Supplementary material for "The Plateau Hemoglobin Paradox: Reversed Effect of Hemoglobin on Surgical Outcomes by Oxygen Saturation Strata at High Altitude": STROCSS 2019 Checklist

### STROCSS 2019 Checklist — 论文 4 (Hb 悖论)

> **稿件版本**: v1.3 (2026-08-18)

> **目标期刊**: International Journal of Surgery (IJS)

> **Checklist 标准**: STROCSS 2019 Guideline (Agha RA, et al. Int J Surg. 2019;72:156-165)

> **填写说明**: 本 md 文件提供章节定位（非页码），党教授后续结合实际 docx 页码填写 PDF 版本

> **参考文献引用**: 稿件[30] = STROCSS 2019 Guideline

---

#### Checklist 填写表

| Item # | Section/Topic | Checklist Item | 稿件章节定位 | Reported? | 备注 |
|---|---|---|---|---|---|
| 1 | Title and abstract | Title and structured abstract (IJS 5 段式) | Title; ABSTRACT (§20-32) | Yes | 摘要 217 词≤250 |
| 2 | Introduction — Background/rationale | Background and rationale | §1 INTRODUCTION (§36-46) | Yes | 高原 Hb 适应与外科应激 |
| 3 | Introduction — Objectives | Objectives and hypotheses | ABSTRACT Objectives; §1 末段 | Yes | Hb 效应在 SpO2 分层反转 |
| 4 | Methods — Study design | Study design | §2.1 Study Design and Setting (§50-53) | Yes | 回顾性单中心队列 |
| 5 | Methods — Setting | Setting, locations, dates, periods of recruitment/follow-up | §2.1 (§50-53) | Yes | 青海红十字医院 2018-2023 |
| 6 | Methods — Participants | Eligibility criteria, sources/methods of selection | §2.2 Participants and Eligibility Criteria (§54-59) | Yes | 612 例成人择期 LC |
| 7 | Methods — Variables | Outcomes, exposures, predictors, potential confounders/effect modifiers | §2.3 Exposure (§60-65); §2.4 Outcome (§66-69); §2.5 Covariates (§70-73) | Yes | Hb/SpO2 暴露；LOS 结局；BMI/年龄/性别/季节 |
| 8 | Methods — Data sources/measurement | For each variable of interest, give sources of data and details of methods of assessment | §2.3-2.5 | Yes | HIS 数据；术前 Hb/SpO2 |

| | | | | | |
|---|---|---|---|---|---|
| 9 | Methods — Bias | Describe any efforts to address potential sources of bias | §2.6 Statistical Analysis (§74-108); §4.5 Limitations (§235-254) | Yes | 多变量回归校正；限制声明 |
| 10 | Methods — Study size | Study size and how it was arrived at | §2.2 (§54-59); §3.1 (§112-131); §3.6 (§185-189) | Yes | 612 例；post-hoc power 0.754 |
| 11 | Methods — Quantitative variables | Explain how quantitative variables were handled | §2.3 Exposure (§60-65); §2.6 Statistical Analysis (§74-108) | Yes | Hb 连续+SpO2 分层 (<93/93-95/≥96) |
| 12 | Methods — Statistical methods | Statistical methods, including confounding and subgroup/interactions | §2.6 Statistical Analysis (§74-108) | Yes | Hb×SpO2 交互；RCS；Bootstrap；LOO |
| 13 | Results — Participants | Numbers at each stage, flow diagram | §3.1 (§112-131); Figure 1 (§277-289) | Yes | 612 例完整流程图 |
| 14 | Results — Descriptive data | Characteristics of participants | §3.1 (§112-131); Table 1 (§423-452) | Yes | 表 1 按 SpO2 分层基线 |
| 15 | Results — Outcome data | Report numbers/outcome events | §3.1-3.2 (§112-151); Table 2 (§453-479) | Yes | LOS 作为连续结局 |
| 16 | Results — Main results | Estimates, confidence intervals, N/A categories | §3.2 Primary Analysis (§132-151); §3.3 Subgroup (§152-163); Table 2-3 | Yes | β=-0.0095, P=.009; mid-SpO2 层 P=.001, d=0.54 |
| 17 | Results — Other analyses | Other analyses (sensitivity, subgroup, mediation) | §3.4 Mediation (§164-178); §3.5 Sensitivity (§179-184); §3.7 Robustness (§191-196) | Yes | 5 项稳健性分析；中介阴性结果诚实报告 |
| 18 | Discussion — Key results | Summarise key results | §4.1 Principal Findings (§199-204) | Yes | SpO2 依赖性 Hb 效应反转 |
| 19 | Discussion — Limitations | Limitations, potential biases, imprecision | §4.5 Limitations (§235-254) | Yes | 8 项限制明确声明 |
| 20 | Discussion — Interpretation | Cautious interpretation | §4.2 Mechanistic (§205-214); §4.3 Clinical Implications (§215-228) | Yes | hypothesis-generating 定位 |
| 21 | Discussion — Generalisability | Generalisability (external validity) | §4.3 Clinical Implications (§215-228); §4.6 Future Directions (§256-266) | Yes | >1.4 亿高原人口；待前瞻复制 |
| 22 | Other information — Funding | Funding sources and role | §9 AUTHORSHIP & FUNDING — Funding (§640 of manuscript MD; typeset page TBD post-production) | Yes | YNZXKT2026009 |
| 23 | Other information — Conflicts of interest | Conflicts of interest | §9 AUTHORSHIP & FUNDING — Conflict of Interest (§643 of manuscript MD; typeset page TBD post-production) | Yes | All authors declare no COI |

#### 党教授后续填写 PDF 指南

1. **下载 STROCSS 2019 Checklist 模板**：

   - URL: https://www.strocss.com/checklist

   - 格式：Word 或 Excel 版

2. **填写"Page/line number"列**：

   - 打开论文 4 docx 文件 (`论文 4_Hb 悖论_JAMA 框架_v1.3_20260818.docx`)

   - 根据 docx 实际页码填写 PDF 中"Page/line in manuscript"列

   - 上表"稿件章节定位"列提供章节对应关系，可作为查找参考

3. **"Reported?"列勾选**：

   - 上表所有项均为"Yes"

   - 如发现某项实际未报告，标记为"Not applicable"并说明

4. **导出 PDF**：

   - 命名：`STROCSS_Checklist.pdf`

   - 存放路径建议：`05_投稿材料/Checklist/`

5. **作为 Supplementary Material 上传**至 IJS 投稿系统

---

#### STROCSS 2019 引用确认

- 稿件中引用位置：§2.1 Study Design and Setting — "STROBE and STROCSS [16,30]"

- 参考文献编号：[30]

- 参考文献条目：Agha RA, Mathou M, Vella-Baldacchino M, Thavayogan R, Orgill RJ; STROCSS Group. STROCSS 2019 Guideline: Strengthening the reporting of cohort studies in surgery. Int J Surg. 2019;72:156-165. doi:10.1016/j.ijsu.2019.11.002

---

本 Checklist 内容由 AI 助手基于稿件章节结构准备，页码字段需党教授结合 docx 实际页码填写 PDF 版本，遵循【坚决杜绝虚构】原则
