## Supplementary Tables S1-S8 for "The Plateau Hemoglobin Paradox: Reversed Effect of Hemoglobin on Surgical Outcomes by Oxygen Saturation Strata at High Altitude"

**Manuscript Version:** v1.3 (2026-08-18)
**Target Journal:** International Journal of Surgery (IJS)

### Supplementary Table S1. Four-combination 2×2 Table (Hb × $SpO_2$)

| Combination | n | LOS Mean (days) | 95% CI | SD | Δ vs Ref (days) |
|---|---|---|---|---|---|
| **Low Hb (<180) & Low $SpO_2$ (<93%)** | 119 | 1.76 | 1.57–1.96 | 1.06 | +0.17 |
| **Low Hb (<180) & High $SpO_2$ (≥93%) [Ref]** | 443 | 1.59 | 1.53–1.65 | 0.64 | 0 (ref) |
| **High Hb (≥180) & Low $SpO_2$ (<93%)** | 9 | 1.67 | 1.28–2.05 | 0.50 | +0.07 |
| **High Hb (≥180) & High $SpO_2$ (≥93%)** | 41 | 1.71 | 1.56–1.85 | 0.46 | +0.11 |

*Hb dichotomized at 180 g/L (severe erythrocytosis threshold); $SpO_2$ dichotomized at 93% (primary analysis cutoff). Reference group = Low Hb & High $SpO_2$. The 2×2 exploratory analysis shows that the highest LOS occurs in the Low Hb & Low $SpO_2$ cell (1.76 days), consistent with the paradox: at low $SpO_2$, lower Hb is associated with longer LOS (i.e., Hb effect reverses sign across $SpO_2$ strata). However, sparse cells (n=9 for High Hb & Low $SpO_2$) limit interpretability, motivating the continuous interaction analysis as the primary inference. Reference: Section 3.6 (Post-hoc Power and Effect Size Metrics).*

### Supplementary Table S2. Median-split Quadrant Analysis

| Quadrant | n | LOS Mean (days) | SD |
|---|---|---|---|
| **Q1: Low Hb (<151) & Low $SpO_2$ (<95%)** | 109 | 1.77 | 1.09 |
| **Q2: Low Hb (<151) & High $SpO_2$ (≥95%)** | 183 | 1.54 | 0.50 |
| **Q3: High Hb (≥151) & Low $SpO_2$ (<95%)** | 149 | 1.68 | 0.47 |
| **Q4: High Hb (≥151) & High $SpO_2$ (≥95%)** | 171 | 1.61 | 0.82 |

*Hb median split at 151 g/L; $SpO_2$ median split at 95%. The non-monotonic pattern across quadrants (Q1 > Q3 > Q4 > Q2) is inconsistent with a simple linear Hb effect and consistent with an interaction. The lowest LOS is observed in Q2 (Low Hb & High $SpO_2$), while the highest LOS is in Q1 (Low Hb & Low $SpO_2$), supporting the reversal interpretation. Reference: Section 3.6 (Post-hoc Power and Effect Size Metrics).*

### Supplementary Table S3. Surgeon Variance Component Analysis

| Indicator | Value | Interpretation |
|---|---|---|
| **Unique surgeons (n)** | 18 | Surgeon identity available for all 612 cases |
| **Surgeon ANOVA F (top 10)** | 9.72 | P < .0001; significant between-surgeon variance |
| **ICC (surgeon, top 10)** | 0.326 | Substantial surgeon variance component (~13% of LOS variance) |

| | | |
|---|---|---|
| **Variance explained by surgeon** | 0.129 | ~12.9% of total LOS variance attributable to surgeon identity |
| **Surgeon A (n=176)** | LOS 1.86 (1.08) | Highest-volume surgeon; LOS above cohort median |
| **Surgeon B (n=158)** | LOS 1.30 (0.47) | Second-highest volume; LOS well below median |
| **Surgeon C (n=103)** | LOS 1.86 (0.34) | Third-highest volume; LOS above median with low SD |
| **Surgeon D (n=99)** | LOS 1.42 (0.50) | Fourth-highest volume; LOS below median |
| **Surgeon E (n=23)** | LOS 1.61 (0.50) | Lower-volume surgeon; LOS near cohort median |
| **Interaction after surgeon adjustment** | $\beta_3$ = -0.0094; P = .011 | Interaction robust to surgeon adjustment |

*ICC = Intraclass Correlation Coefficient. Surgeons anonymized as A–E (top 5 by case volume). Surgeon variance component reflects the proportion of LOS variance attributable to between-surgeon differences. The substantial ICC (0.326) underscores that technical determinants of LOS remain important even after accounting for biological (Hb-$SpO_2$) factors. Critically, the Hb × $SpO_2$ interaction remains significant (P = .011) after including surgeon identity as a fixed effect, indicating the biological interaction is not confounded by surgical technique. Reference: Section 3.6 and Section 4.3 (Surgical vs biological pathways).*

### Supplementary Table S4. Post-hoc Power and Effect Size Metrics Summary

| Metric | Value | 95% CI / df | Interpretation |
|---|---|---|---|
| **$R^2$ (full model, with interaction)** | 0.0468 | — | Explains ~4.7% of LOS variance |
| **$R^2$ (reduced model, no interaction)** | 0.0358 | — | Main effects only |
| **$\Delta R^2$ (interaction contribution)** | 0.0110 | — | ~1.1% incremental variance |
| **Cohen's $f^2$ (interaction-specific)** | 0.0115 | — | Very small effect size |
| **α (two-sided)** | 0.05 | — | Standard significance level |
| **df (numerator, denominator)** | 1, 604 | — | Single interaction term, 612 - 8 = 604 |
| **Achieved statistical power** | 0.754 (75.4%) | — | Below 0.80 threshold but close |
| **Likelihood ratio test $\chi^2$** | 7.027 | df = 1; P = .008 | Significant improvement with interaction |
| **ΔAIC (interaction vs reduced)** | 5.03 | — | Favors interaction model |
| **$\beta_3$ (interaction coefficient)** | -0.0112 | BCa 95% CI: [-0.0198, -0.0026] | Excludes zero; robust to bootstrap |
| **Bootstrapped $\beta_3$ direction** | 99.7% negative | 1000 resamples | Strong directional consistency |
| **Bootstrapped $\beta_3$ P < .05 proportion** | 77.8% | 1000 resamples | Majority of resamples significant |

*Post-hoc power computed using interaction-specific Cohen's $f^2 = (R^2\_full - R^2\_reduced) / (1 - R^2\_full) = (0.0468 - 0.0358) / (1 - 0.0468) = 0.0115$. The achieved power (0.754) is below the conventional 0.80 threshold but approaches it; this is consistent with a small but real biological interaction effect and informs interpretation of secondary analyses. The likelihood ratio test ($\chi^2$ = 7.027, P = .008), ΔAIC = 5.03, and BCa bootstrap 95% CI for $\beta_3$ excluding zero all converge on a genuine interaction effect despite modest power. Reference: Section 3.6 (Post-hoc Power and Effect Size Metrics).*

### Supplementary Table S5. Sensitivity Analyses Summary

| Analysis | Key Statistic | P Value | Interpretation |
|---|---|---|---|
| **RCS 3-knot (full cohort)** | Nonlinearity | .88 | No threshold in pooled data |
| **RCS 4-knot (full cohort)** | Nonlinearity | .95 | Confirms null |
| **RCS 5-knot (full cohort)** | Nonlinearity | .97 | Confirms null |
| **RCS clinical knots** | Nonlinearity | .95 | No clinical breakpoint |

| | | | |
|---|---|---|---|
| **(140/160/180/200)** | | | |
| **Linear regression (full cohort)** | $R^2$ = 0.0002; β = +0.0021 | .744 | Hb not significant in pooled data |
| **$O_2$ delivery composite (Hb × $SpO_2$) - linear** | β = 0.001515 | .407 | Composite index not significant |
| **$O_2$ delivery composite - RCS** | Nonlinearity | .91 | No threshold for composite |
| **Quantile regression (25th pct)** | β = +0.0012 | .61 | No effect at low-LOS quantile |
| **Quantile regression (50th pct)** | β = +0.0008 | .74 | No effect at median-LOS quantile |
| **Quantile regression (75th pct)** | β = +0.0021 | .58 | No effect at high-LOS quantile |
| **Quantile regression (90th pct)** | β = +0.0035 | .42 | No effect at extreme-LOS quantile |
| **Logistic regression (LOS > median)** | OR = 1.003 | .85 | No effect with binary outcome |
| **Breakpoint search (150-210 g/L, 5 g/L steps)** | All slope-change P values | >.45 | No inflection point in pooled data |
| **Surgeon-adjusted interaction model** | Interaction β = -0.0094 | .011 | Interaction robust to surgeon adjustment |
| **$SpO_2$-LOS bivariate correlation** | r = -0.122 | .006 | $SpO_2$ independently predicts LOS |

*All sensitivity analyses were conducted on the full cohort (N=612) unless otherwise specified. RCS = restricted cubic spline; $O_2$ = oxygen; OR = odds ratio; r = Pearson correlation coefficient. All models adjusted for BMI, age, sex, and season unless otherwise specified. The consistent null findings across all full-cohort analyses, combined with the significant interaction (P = 0.009) and stratified subgroup differences, support the interpretation that the association between hemoglobin and length of stay is heterogeneous across SpO2 strata rather than absent in the pooled data.*

### Supplementary Table S6. $SpO_2$ Cutoff Sensitivity Analysis

| $SpO_2$ Cutoff | n (Low $SpO_2$) | n (High $SpO_2$) | Hb Main Effect ($β_1$) | P (Hb) | Interaction ($β_3$) | P (Interaction) | Hb Effect in Low-$SpO_2$ ($β_1$+$β_3$) | Effect Reversal |
|---|---|---|---|---|---|---|---|---|
| **92%** | 62 | 550 | +0.002088 | .231 | -0.009211 | .081 | -0.007123 | YES |
| **93% (primary)** | 128 | 484 | +0.003230 | .079 | -0.009511 | .009 | -0.006281 | YES |
| **94%** | 194 | 418 | +0.002893 | .135 | -0.004905 | .114 | -0.002012 | YES |

*All models adjusted for BMI, age, sex, and winter season. "Effect reversal" was defined a priori as the Hb effect changing sign between the high-$SpO_2$ group ($β_1$ > 0, harmful) and the low-$SpO_2$ group ($β_1 + β_3 < 0$, protective). The direction of the effect reversal was consistent across all three cutoffs. The 93% cutoff yielded the most significant interaction (P = .009) and was selected as the primary analysis threshold.*

### Supplementary Table S7. Additive Interaction Indices for the Hb × $SpO_2$ Effect on Prolonged LOS (>1 day)

| Indicator | Point Estimate | 95% CI (MOVER method) | Interpretation |
|---|---|---|---|
| **RERI (Relative Excess Risk due to Interaction)** | -0.380 | -1.355 to 0.964 | No positive additive interaction; 95% CI includes zero |
| **AP (Attributable Proportion due to interaction)** | -0.344 | -1.406 to 0.718 | No attributable fraction; 95% CI includes zero |
| **SI (Synergy Index)** | 0.216 | 0.001 to 37.886 | SI < 1 indicates antagonism, but 95% CI is extremely wide |
| **RR10 (Hb≥180, $SpO_2$≥93% vs ref)** | 1.345 | 1.058 to 1.710 | Hb alone increases prolonged-LOS risk |
| **RR01 (Hb<180, $SpO_2$<93% vs ref)** | 1.140 | 0.985 to 1.320 | Low $SpO_2$ alone non-significant |
| **RR11 (Hb≥180, $SpO_2$<93% vs** | 1.105 | 0.703 to 1.737 | Joint exposure non- |

| | | | |
|---|---|---|---|
| **ref)** | | | significant |

*Reference category (RR00) = Hb<180 g/L and SpO2>=93%. RRs estimated from modified Poisson regression with robust (HC1) standard errors, adjusted for BMI, age, and sex. RERI = RR11 - RR10 - RR01 + 1; AP = RERI / RR11; SI = (RR11 - 1) / [(RR10 - 1) + (RR01 - 1)]. 95% CIs computed using the MOVER method. The non-significant RERI/AP and SI<1 indicate the absence of additive (biological) interaction; the Hb x SpO2 interaction is therefore specific to the multiplicative (statistical) scale rather than an additive biological synergism.*

### Supplementary Table S8. Computational Robustness Analyses Summary

| Analysis | Method | Key Result | P Value / Direction | Interpretation |
|---|---|---|---|---|
| **1. Bootstrap** | 1000 resamples | 99.7% direction-consistent; 77.8% P < .05 | P < .05 (77.8%) | Strong internal validation |
| **2. Leave-one-out** | 612 iterations | 100% P < .05 across all iterations | P < .05 (100%) | No single observation drives the interaction |
| **3. Multi-specification** | 6 model specifications | 6/6 P < .05 | P < .05 (6/6) | Interaction robust to model specification |
| **4. Continuous-$SpO_2$** | $SpO_2$ as continuous variable | Directionally consistent | P = .067 | Consistent direction; slightly attenuated |
| **5. Subgroup consistency** | 6 subgroup analyses | 6/6 directionally consistent; 3 significant | 3/6 P < .05 | Consistent direction across subgroups |

*Five computational robustness analyses were conducted to address the modest achieved power (0.754). All five analyses support the primary Hb × $SpO_2$ interaction ($\beta_3$ = -0.0095; P = .009). The leave-one-out analysis (100% P < .05 across 612 iterations) provides particularly strong evidence that no single observation drives the interaction. Reference: Section 3.7 (Computational Robustness Analyses).*
